# LLM-assisted evidence audit of late-stage cancer incidence as a screening trial endpoint

**DOI:** 10.64898/2026.08.29.26361733

**Authors:** Shiyin Li, Wanheng Zhang, Xing Xing, Ziyuan Shen, Yining Wang, Zhao Chen, Onicio Batista Leal Neto, Yulin Yu, Chong Wu, Lifeng Lin

## Abstract

**Background:** Late-stage cancer incidence is being considered as an earlier endpoint in cancer-screening trials, but its trial-level association with cancer-specific mortality may depend on evidence selection and endpoint harmonization. We evaluated the robustness of this association to source-verified additions.

**Methods:** We reconstructed the PubMed corpus underlying a 41-comparison review. Gemini 3.1 Pro Preview was used only to prioritize reports for blinded human reassessment. Reviewers determined eligibility, linked reports from the same trial, harmonized endpoints, and verified comparison-level data. We recalculated unweighted Pearson correlations overall and by cancer type after adding earliest-compatible trial comparisons.

**Results:** Among 1209 candidate records, 996 PDFs were assessed. Thirty-three reports absent from the source review were prioritized; 26 were eligible, representing 18 trials, and 8 provided compatible comparisons. Adding these comparisons increased the dataset from 41 to 49 and attenuated the overall correlation from 0.73 (95% confidence interval [CI] = 0.55 to 0.85) to 0.59 (95% CI = 0.37 to 0.75). Updated correlations were 0.49 (95% CI = -0.26 to 0.87) for breast, - 0.23 (95% CI = -0.71 to 0.40) for colorectal, and 0.83 (95% CI = 0.54 to 0.95) for lung cancer. One sparse-event comparison influenced the colorectal estimate.

**Conclusions:** The overall association was sensitive to evidence composition, and cancer-specific stability varied. Late-stage incidence should be evaluated by cancer type and with prespecified sensitivity analyses for evidence selection and endpoint definitions. Model-assisted prioritization cannot replace human eligibility review, trial reconciliation, and source verification.

## 1. Introduction

Cancer-specific mortality is the definitive outcome for evaluating the benefit of cancer screening. However, mortality endpoints often require large cohorts and prolonged follow-up, during which screening tests and cancer treatments may change.[1] Late-stage cancer incidence is therefore being considered as an earlier endpoint, although its use requires assumptions about disease progression and screening effects.[2] The UK National Screening Committee regards the incidence of stage III-IV cancer as a promising early indicator but not a replacement for mortality,[3] because evidence is strongly dependent on cancer type. In a systematic review and meta-analysis of randomized cancer-screening trials in *JAMA* by Feng et al.,[4] the overall Pearson correlation coefficient was 0.73 between the incidence of stage III-IV cancer and cancer-specific mortality, but cancer-specific correlations ranged from 0.99 for ovarian, 0.92 for lung, 0.70 for breast, 0.39 for colorectal, and -0.68 for prostate cancer in their analysis of 41 comparisons. Rebolj et al. later reported an overall correlation of 0.69 across 61 comparisons, with estimates of 0.91 for lung, 0.79 for breast, and 0.58 for colorectal cancer.[5] These differences suggest that conclusions may depend on cancer type and on the reports, follow-up periods, stage definitions, and comparisons that finally enter the synthesis after screening.

Evidence recovery is therefore part of the surrogate-endpoint question, not merely an administrative step in review production. Trial evidence may be distributed across primary publications, extended follow-up publications, subgroup analyses, appendices, and supplements. A potentially relevant publication may still be unsuitable for synthesis until reviewers reconcile trial identity, follow-up, stage definitions, denominators, and duplicate comparisons. Kirkham et al.[6] found that adjustment for missing primary-outcome data changed treatment estimates by at least 20% in 18 of 81 reviews and rendered 8 of 42 significant findings nonsignificant. Tramèr et al.[7] found that duplicated data overestimated efficacy by 23%. These examples show how incomplete or duplicated evidence can alter quantitative conclusions. Large language models (LLMs) may help prioritize full texts for reassessment, but their outputs vary by model, prompt, corpus, and task and require source-level human verification.[8, 9] Cao et al.[10] found that optimized prompts achieved weighted sensitivities of 97.7% and 96.5% for abstract and full-text screening, whereas zero-shot prompts achieved approximately 49% at both stages. Dennstädt et al.[11] found sensitivity ranged from 81.9% to 97.6%, and specificity ranged from 19.1% to 75.2%, with changes after minor prompt revisions. Other studies have addressed human-in-the-loop data extraction,[12] structural failures in meta-analysis evidence extraction,[13] and agreement with systematic reviews’ conclusions.[14] Evidence-synthesis extraction studies also show that performance varies by model, tool chain, and data-field complexity, reinforcing the need for source-level human verification.[15, 16] These studies mainly evaluated agreement with existing labels rather than the downstream validity of model-positive publications absent from the source-review included-publication set.

In this study, we evaluated LLMs as an end-to-end, human-supervised evidence audit. Using the 41-comparison primary framework by Feng et al.,[4] we examined whether repeated full-text LLM assessments could identify a compact set of publications for human reassessment. This design is related to recent full-agreement multi-LLM screening strategies, but extends the evaluation from citation triage to full-text evidence auditing and assessment of downstream synthesis stability.[17] We then traced additional publications through human eligibility assessment, linking multiple publications of the same underlying randomized trial, human endpoint verification, and quantitative inclusion. Finally, we tested whether reviewer-verified additions changed cancer-specific stage and mortality endpoints. The study therefore evaluates the stability of an evidence synthesis after audited additions, not autonomous screening or definitive surrogate validation.

## 2. Methods

### 2.1 Study design

We conducted a retrospective evidence audit of the cancer-screening review by Feng et al.[4] The LLM prioritized full-text reports for human reassessment; publication-level agreement was assessed against the inherited reference set, and quantitative analyses were performed at the intervention-control comparison level after trial-family reconciliation. The primary quantitative outcome was the change in cancer-specific correlation between reductions in cancer-specific mortality and incidence of stage III-IV cancer after adding reviewer-verified comparisons. No individual participant data were used. This retrospective audit was not prospectively registered, and no study-specific protocol was deposited before the analyses.

The candidate corpus comprised 1,209 PubMed-indexed publications identified by reproducing Feng et al.’s original search strategy with the same date restriction; the search period was not extended. The source review contained 60 publications, of which 58 were present in the audit corpus and formed the reference set. Feng et al.’s primary stage III-IV analysis used 39 publications contributing 41 intervention and comparison groups. A separate dataset of 63 comparisons retained multiple follow-up observations from some trials (Supplementary Table S7). Because the 58 reference-set publications were not independently re-adjudicated, reference-set recovery measured agreement with inherited decisions rather than true screening sensitivity or accuracy.

The original PubMed NBIB file was imported into Zotero. PDFs were retrieved automatically and through manual searches, then matched to the corpus by PMID. Full-text PDFs were available for 996 records; the remaining 213 were not assessed or classified as negative. The assessed publications spanned 1977-2024 (median, 2014; interquartile range, 2007-2018).

### 2.2 Full-text LLM audit and consensus classification

The fixed prompt asked whether a full-text report warranted human reassessment and directed the model to inspect the main text, tables, embedded appendices, and references to separate supplements before exclusion. This full-text emphasis is consistent with evidence-synthesis NLP (natural language processing) work showing that systematic literature review data extraction often requires information beyond titles and abstracts.[18] An LLM inclusion decision indicated either that required data were located in the report or in supplementary material. Cancer-specific mapping rules, adapted from Feng et al., covered breast tumor size, colorectal Dukes stage, lung unresectability, and prostate stage or risk categories. Supplementary Appendix S1 provides the complete prompt and its output categories.

Gemini 3.1 Pro Preview was selected because, at the time of the audit, it supported native PDF input, a 1,048,576-token context window, multimodal document understanding, structured output, and API-level parameter control, capabilities suited to long clinical reports containing tables and figures.[19] PDFs were submitted through the native PDF route without local text extraction or optical character recognition. Each PDF underwent five repeated assessments using the Google Gemini 3.1 Pro Preview model via OpenRouter’s application programming interface with temperature 1.0, which advised by Gemini 3 [19]; the distribution of the resulting five-run verdict patterns is reported in Supplementary Table S1. This repeated evaluation has also been used by Oami et al.[20] in LLM-assisted screening, the same prompt and settings were used throughout. Technical implementation, sampling defaults, ordering, and error handling are described in Supplementary Appendix S2.

For each publication, we counted the number of LLM inclusion decisions across the five repetitions. Under the post hoc five-of-five rule, only reports with inclusion-type decisions in all five assessments were prioritized; any error therefore prevented prioritization. The rule was not considered prespecified, optimal, or externally validated, and the repetitions were not treated as independent validators. We also examined thresholds requiring at least one, two, three, or four LLM inclusion decisions. Complete human assessment was available only for publications absent from the reference set and prioritized by the 5-of-5 rule.

At each threshold, we reported the total number of the 996 PDF-assessed publications meeting the rule. Reference-set recovery was the number of reference-set publications meeting the rule. Potential workload reduction was calculated as the number and percentage of those 938 publications not meeting the rule. These quantities describe agreement with an inherited reference set, not true screening performance.

### 2.3 Human adjudication and endpoint reconciliation

The 5-of-5 rule prioritized 91 publications: 58 in the reference set and 33 additional reports (Supplementary Table S2). Two reviewers (ZS and YW) whose institutional backgrounds were in clinical and medical research independently assessed publication-level eligibility for all 33 publications using a form that omitted model verdicts and previous human decisions. Three disagreements underwent final adjudication. Consequently, 26 publications were retained at the publication level, and 7 were excluded (Supplementary Table S3); the retained reports were linked into 18 unique trials for endpoint reconciliation.

The 905 publications outside the 5-of-5 set, 213 records without mapped PDFs, and 58 reference-set publications were not re-adjudicated; therefore, eligibility among nonprioritized or unavailable records could not be estimated.[21] For each of the 26 retained publications, we compared four provisional arm-level event counts across the five assessments, including cancer-specific deaths and late-stage cancers in both the intervention and control arms. No model-derived count entered the sensitivity analysis without manual verification against source publications.

### 2.4 Unique trials consolidation

Consistent with Cochrane guidance, we collated publications from the same trial so that the trial, rather than the publication, was the unit of interest.[22] The 26 retained publications represented 18 unique trials (Supplementary Table S4). For each trial, we documented cancer type, follow-up, endpoint definition, denominators, selected publication, and overlap with the source review. We excluded duplicate papers for the same trial and publications with incompatible denominators, subgroup-only results, model-derived estimates, or missing endpoint counts. Eight unique trials supplied compatible comparison-level data. For each recovered trial contributing to the update, we selected the earliest follow-up reporting compatible mortality and late-stage or trial-specific proxy outcomes for the same randomized population, approximating Feng et al.’s earliest- follow-up framework.

Two additional reviewers independently verified the eight intervention-control comparisons against the source publications (Supplementary Table S5). They checked denominators, event counts, arm assignment, and analysis population.

### 2.5 Sensitivity analysis

We used Feng et al.’s primary stage III-IV dataset of 41 intervention and comparison groups as the baseline. Adding eight reviewer-verified intervention-control comparisons, one from each recovered trial, produced a 49-comparison dataset (Supplementary Table S6). We did not update the separate dataset of 63 follow-up comparisons because it contained repeated observations from some trials.

For each endpoint, percentage reduction was calculated as (control-arm risk - intervention-arm risk)/control-arm risk. Positive values indicated lower endpoint risk in the intervention arm, whereas negative values indicated higher risk. We excluded records with zero control-arm risk or irreconcilable counts, denominators, analysis populations, or trial relationships.

We calculated descriptive, unweighted Pearson correlation coefficients (*r*) for the original 41 and updated 49 comparisons. Analyses were conducted overall and separately for breast, colorectal, lung, ovarian, and prostate cancer. The least-squares regressions used mortality reduction as the dependent variable and late-stage reduction as the independent variable. We fitted both ordinary least-squares models with an intercept and no-intercept models constrained through the origin. Remaining cancer types were combined descriptively and were not interpreted as one cancer-specific stratum. Comparisons were not weighted by sample size, event count, or statistical precision.

We calculated 95% confidence intervals for Pearson *r* values using Fisher’s z transformation.[23] For a descriptive summary of variation among cancer types, we transformed the five principal cancer-specific correlations to Fisher z values. Each transformed correlation was weighted by n-3. We calculated the Q and I² statistics.[24, 25] These weights were used only for the between-cancer summary.

For the original 41 comparisons, P<.05 classifications were transcribed from Feng et al.’s Supplementary eTable 2. For the eight additions, two-sided chi-square tests with continuity correction compared event proportions; a reduction was classified as significant only when it was positive and (P<.05). Each comparison was assigned to one of four mutually exclusive categories: significant reductions in both endpoints, neither endpoint, late-stage incidence only, or mortality only. The first two categories were considered concordant and the latter two discordant.

The update was restricted to the incidence of stage III-IV cancer and the 41-comparison primary baseline. We did not re-estimate analyses using P<0.10, stage IV alone, stage shift, last or all follow-up periods, or colorectal screening modality. These analyses required verified variables or follow-up reconciliation rules that were unavailable.

## 3. Results

### 3.1 Audit corpus and consensus performance

Of 1,209 records, 996 (82.4%) had mapped PDFs and underwent five repeated assessments, whereas 213 were unassessed and not classified as negative. Of the 60 source-review publications, 58 were present in the corpus and formed the reference set. Alternative consensus thresholds and complete five-run verdict distributions are reported in Supplementary Table S1, S2, and S8. Under the 5-of-5 rule, 91 publications were prioritized, comprising all 58 reference-set publications and 33 publications absent from the reference set (Figure 1, Supplementary Tables S1 and S2).

**Figure 1.**
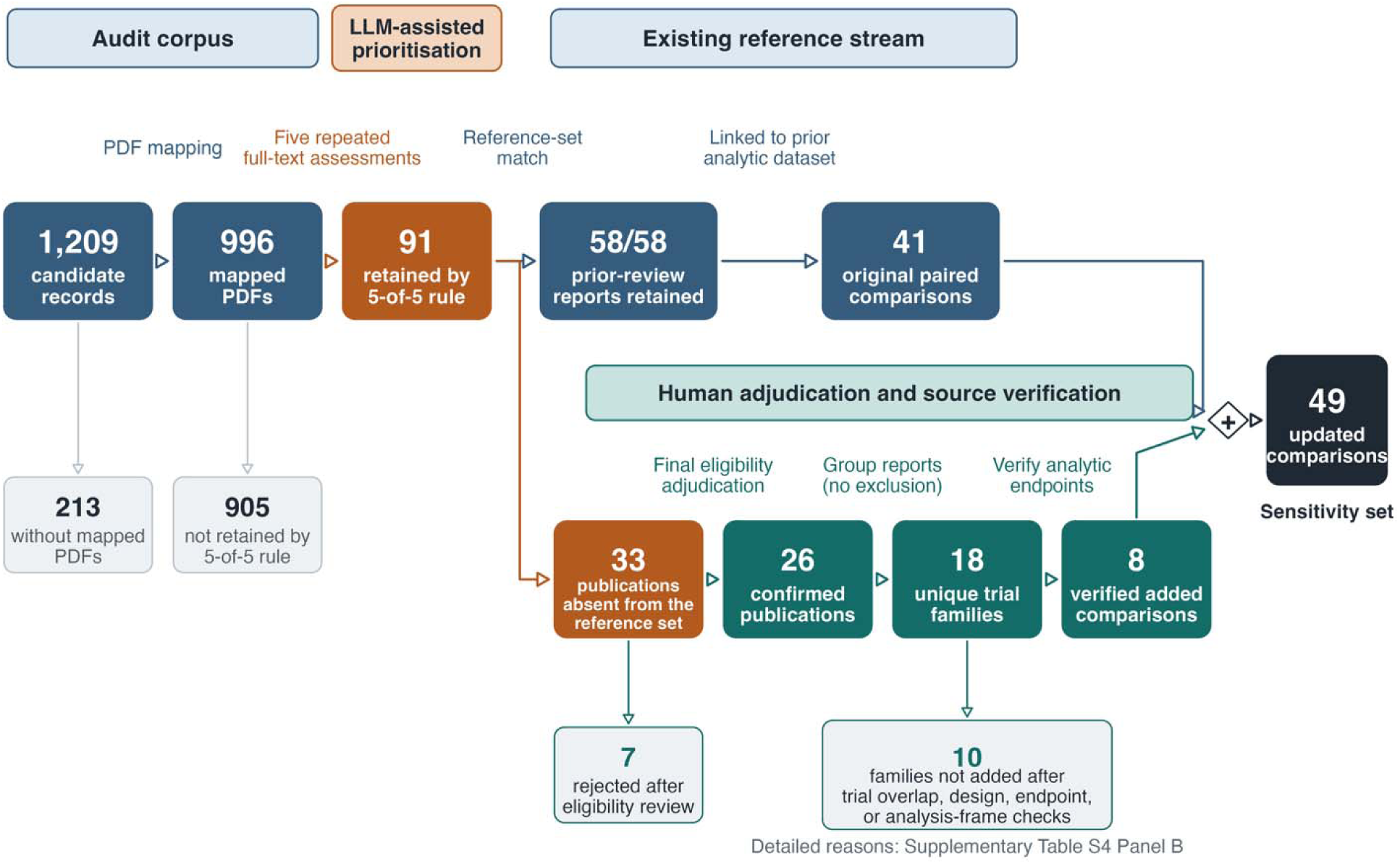
Human-supervised evidence-audit pathway.

Only the 33 prioritized nonreference reports underwent human reassessment; the 905 nonprioritized reports, 213 records without mapped PDFs, and 58 reference-set reports were not re-adjudicated, so true accuracy and recall could not be estimated. Two masked reviewers agreed on 30 of the 33 additional reports (90.9%; Cohen’s κ = 0.68); after adjudication, 26 of 33 (78.8%; 95% CI, 62.2% to 89.3%) were eligible (Table 1; Supplementary Table S3).

**Table 1.** Human review and endpoint-data availability among publications retained but absent from the included publications from Feng et al.

| Step | n | PMID(s) |
| --- | --- | --- |
| Sent to human reassessment | 33 | 3411625[35], 2404878[36], 2035504[37], 7579497[38], 7736395[39], 9386870[40], 1290631[41], 8340943[42], 8898422[43], 10365903[44], 11147613[45], 19339719[46], 23065684[47], 19637354[48], 25698407[49], 29224236[50], 31221620[51], 31537697[52], 1854979[53], 7497137[54], 23750991[55], 33141657[56], 26095467[57], 28976536[58], 30653262[59], 34817559[60], 37657461[61], 11147611[62], 25372087[63], 31336289[64], 25556937[65], 37029074[66], 25777669[67] |
| Confirmed eligible | 26 | 3411625, 2404878, 2035504, 7579497, 7736395, 9386870, 1290631, 8898422, 10365903, 11147613, 19339719, 23065684, 19637354, 25698407, 29224236, 31221620, 31537697, 1854979, 7497137, 23750991, 33141657, 26095467, 28976536, 30653262, 34817559, 37657461 |
| Rejected | 7 | 11147611, 25372087, 31336289, 25556937, 37029074, 25777669, 8340943 |
| Rejected: endpoint not usable | 5 | 11147611, 25372087, 31336289, 25556937, 37029074 |
| Rejected: non-random allocation | 1 | 8340943 |
| Rejected: language | 1 | 25777669 |
| Stable extracted endpoint-count set | 17 | 3411625, 2404878, 2035504, 7579497, 7736395, 9386870, 1290631, 8898422, 10365903, 11147613, 19339719, 23065684, 19637354, 25698407, 29224236, 31221620, 31537697 |
| Manually resolved extracted endpoint-count set | 4 | 1854979, 7497137, 23750991, 33141657 |
| Not suitable for quantitative inclusion | 5 | 26095467, 28976536, 30653262, 34817559, 37657461 |
| Reconciled unique trials | 18 | 26 confirmed-eligible PMIDs grouped into 18 unique trials |
| Analyzable unique trials records | 8 | 7579497, 7497137, 33141657, 19339719, 8898422, 23750991, 2404878, 31537697 |

### 3.2 Sensitivity analysis of the stage-mortality association

The sensitivity analysis used Feng et al.’s 41 primary paired comparisons as its baseline. Adding eight reviewer-verified unique trial-level comparisons produced a 49-comparison dataset. Across all cancer types, the unweighted Pearson correlation decreased from *r* = 0.73 (95% CI, 0.55 to 0.85) to *r* = 0.59 (95% CI, 0.37 to 0.75) (Table 4, Figure 2). The corresponding regression slope decreased from 0.66 to 0.52. Descriptive I² across the five principal cancer-specific correlations increased from 64.7% to 74.9%.

**Figure 2.**
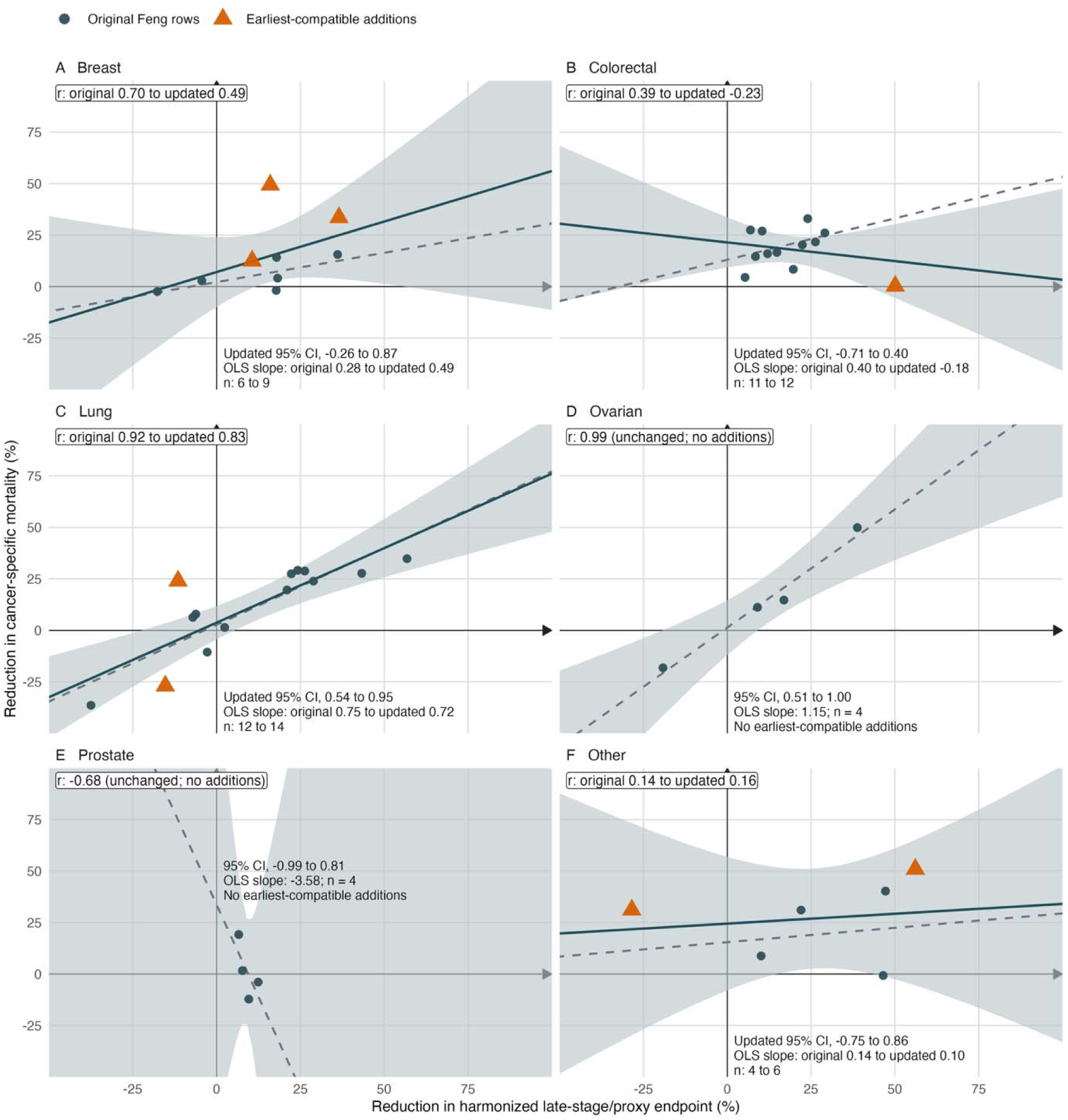
Cancer-type-specific stage-mortality associations before and after adding eight endpoint-verified comparisons. Each point represents one intervention-control comparison from the complete 49-comparison dataset (Supplementary Table S6). Filled circles show the 41 original comparisons from Feng et al., and orange triangles show the added comparisons. Dashed lines show the original unweighted regression fits, whereas solid lines show the updated fits.

For cancer-specific correlations, adding three breast comparisons changed Pearson r from 0.70 (95% CI, -0.26 to 0.96) to 0.49 (updated 95% CI, -0.26 to 0.87). Adding one colorectal comparison changed r from 0.39 (95% CI, -0.27 to 0.80) to -0.23 (updated 95% CI, -0.71 to 0.40). Adding two lung comparisons changed r from 0.92 (95% CI, 0.72 to 0.98) to 0.83 (0.54 to 0.95). The colorectal update was influenced by one sparse comparison of 400 intervention and 399 control participants, with one cancer-specific death in each arm and one versus two proxy late-stage events. Ovarian and prostate estimates were unchanged because no comparisons were added to those strata. In the heterogeneous group of other cancers, *r* changed from 0.14 to 0.16 and was reported only descriptively.

Significance classifications were concordant for 29 of 41 original comparisons (70.7%) and 36 of 49 updated comparisons (73.5%). However, five of the seven concordant additions were nonsignificant for both endpoints; discordance increased from 12 to 13 because the Stockholm Mammography Trial showed a mortality-only reduction (Table 5).

### 3.3 Endpoint reconciliation and unique-trial consolidation

Repeated LLM assessments returned the same four arm-level endpoint counts for 17 of 26 eligible publications (Supplementary Table S4). This agreement indicated repeatability, not verified accuracy [26]. Manual source review resolved discrepancies. Five publications lacked compatible randomized-arm data because exact counts were unavailable, because only adjusted or modeled estimates were reported, or because the analyses were restricted to incompatible subgroups or case-mortality populations (Table 1).

The 26 retained publications represented 18 unique trials (Table 2; Supplementary Table S4); eight yielded compatible, reviewer-verified comparisons. Of the remaining 10 trials, seven overlapped with trials already represented in Feng et al. Three lacked data compatible with the target analysis. The eight added comparisons comprised three breast, two lung, one colorectal, one cervical, and one liver cancer comparison (Table 3). Two reviewers verified all eight comparisons; their review corrected the Swedish Two-County mortality counts from 269/277 to age-compatible counts of 224/238, while the stage counts remained 389/432 (Supplementary Table S5).

**Table 2.** Unique trials reconciliation of the 26 publications retained after human full-text reassessment.

| Unique trial | Cancer type | Feng et al.'s included record | LLM-retained, human-confirmed PMID | Selected PMID | Rationale |
| --- | --- | --- | --- | --- | --- |
| *Czech Lung Screening Trial* | Lung |  | 11147613, 2404878 | 2404878 | Earliest-compatible follow-up |
| *ERSPC Rotterdam treatment-difference analysis* | Prostate | 23759326 | 25698407 | NA | Same unique trials as Feng et al-included records |
| *FaMRisc Trial* | Breast |  | 31221620 | NA | zero breast-cancer deaths and deferred mortality reporting. |
| *Finnish Randomized Study of Prostate Cancer Screening* | Prostate | 23479454 | 29224236, 30653262, 34817559 | NA | Same unique trials as Feng et al-included records |
| *HIP Breast Screening Trial* | Breast |  | 3411625, 2035504 | NA | Not chosen. Reported case-mortality among women diagnosed with breast cancer, not full randomized-arm breast-cancer mortality. |
| *Johns Hopkins Lung Project / Memorial Sloan-Kettering Lung Study combined sputum-cytology analysis* | Lung | 6734291 | 19637354 | NA | Same unique trials as Feng et al-included records |
| *LungSEARCH Trial* | Lung |  | 31537697 | 31537697 |  |
| *PLCO Flexible Sigmoidoscopy Trial* | Colorectal | 22612596, 30502933 | 28976536 | NA | Same unique trials as Feng et al-included records |
| *PLCO ROCA ovarian-screening analysis* | Ovarian | 21642681 | 23065684 | NA | Same unique trials as Feng et al-included records |
| *Randomized hepatocellular carcinoma surveillance trial* | Liver / hepatocellular carcinoma |  | 23750991 | 23750991 |  |
| *Rural India HPV Screening Trial* | Cervical |  | 19339719 | 19339719 |  |
| *Stockholm Mammography Trial* | Breast |  | 1854979, 7579497, 9386870 | 7579497 | Earliest-compatible follow-up |
| *Swedish / Norrköping prostate DRE screening trial* | Prostate | 15548438, 21454449 | 1290631 | NA | Same unique trials as Feng et al-included records |
| *Swedish Two-County Trial* | Breast |  | 7497137, 7736395 | 7497137 | Earliest-compatible follow-up |
| *Telemark Polyp Study I* | Colorectal |  | 8898422, 10365903 | 8898422 | Earliest-compatible follow-up |
| *UK Age RCT* | Breast |  | 33141657 | 33141657 |  |
| *UKCTOCS exploratory high-grade serous analysis* | Ovarian / tubo-ovarian | 26707054, 33991479 | 37657461 | NA | Same unique trials as Feng et al-included records |
| *Unprovoked VTE occult-cancer screening trial* | Multiple / occult cancers |  | 26095467 | NA | not suitable for quantitative inclusion |

**Table 3.**
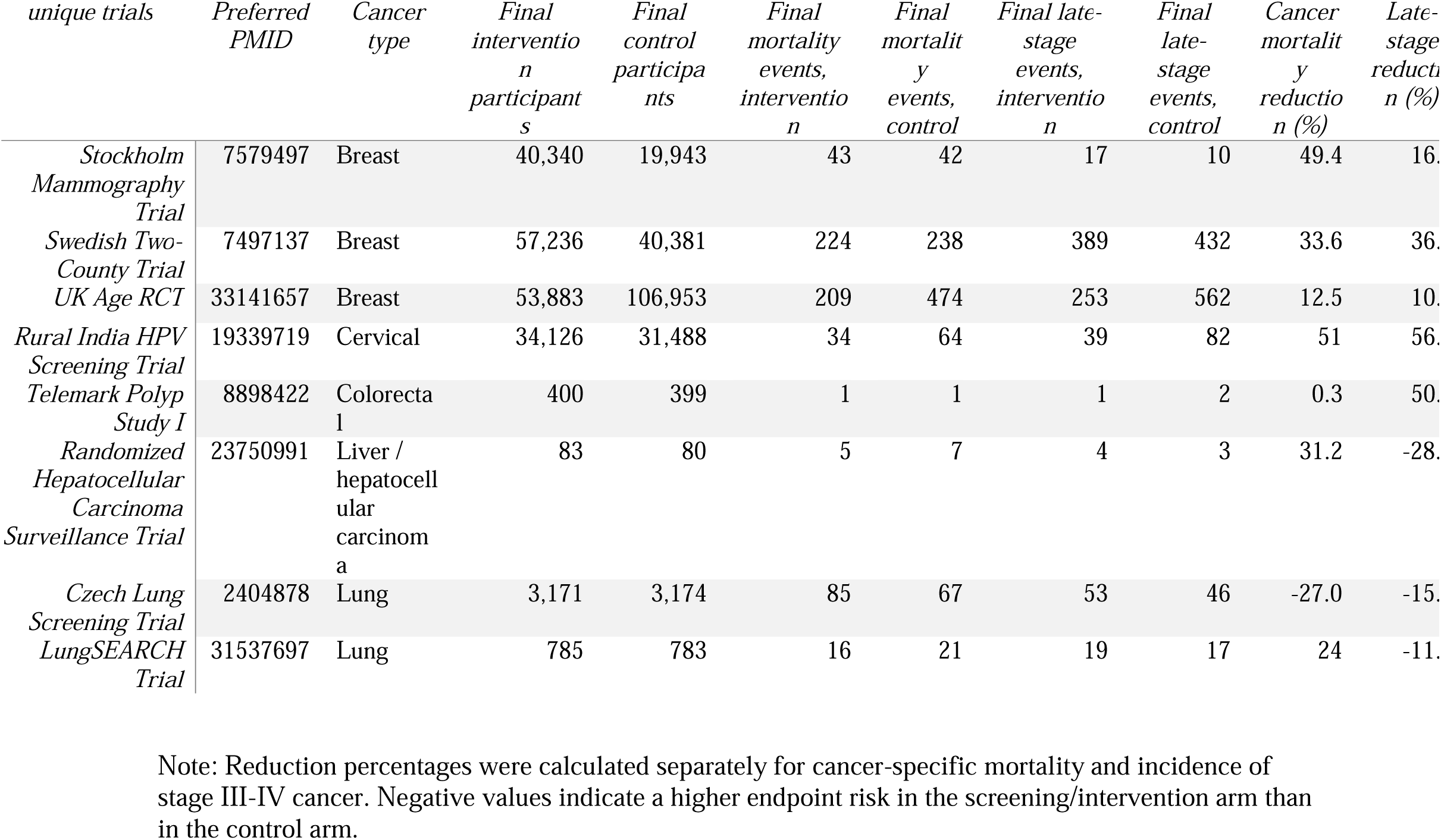
Final endpoint data for the eight comparisons included in the sensitivity analysis.

**Table 4.**
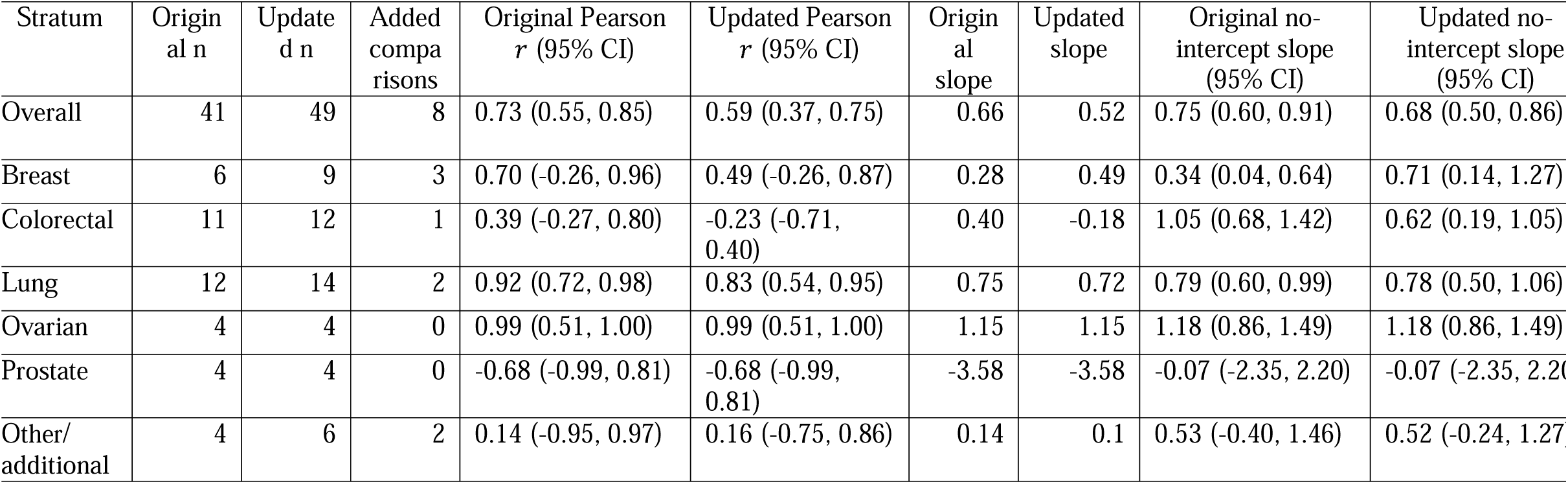
Stage-mortality associations before and after adding eight endpoint-verified comparisons, overall and by cancer type.

**Table 5.**
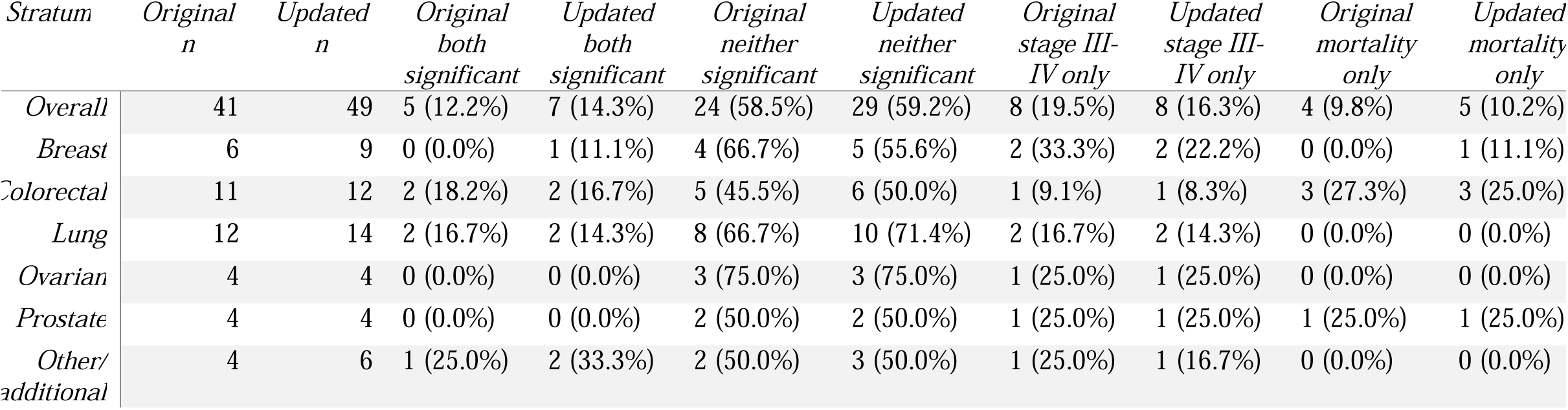
Concordance of statistical significance for mortality and stage III-IV incidence before and after the sensitivity update (two-sided α = 0.05)

## 4. Discussion

This evidence audit showed that recovered reports did not translate directly into independent quantitative evidence, but source-verified additions changed the estimated association between late-stage incidence and cancer-specific mortality. Of 33 prioritized nonreference reports, 26 were eligible, representing 18 trials and eight compatible comparisons. Adding these comparisons reduced the overall correlation from 0.73 to 0.59 and increased descriptive heterogeneity across the five principal cancer types from 64.7% to 74.9%. The association remained strong for lung cancer, became weaker and uncertain for breast cancer, and was inconclusive for colorectal cancer; ovarian and prostate estimates were unchanged because no comparisons were added. Although binary significance concordance increased, most concordant additions were non-significant for both endpoints, so the higher percentage did not indicate that endpoint discordance had been resolved. Neither correlation nor binary significance concordance establishes that an intervention’s effect on an earlier endpoint reliably predicts its effect on mortality.[27, 28]

These findings have implications for cancer-screening trial design. Late-stage incidence may shorten follow-up and improve feasibility, but one overall correlation can obscure substantial cancer-specific variation and sensitivity to evidence composition. Our update preserves the broad conclusion of Feng et al. and the UK National Screening Committee that late-stage incidence may be informative for some cancers but should not replace mortality in definitive evaluations.[3] The less stable breast and colorectal estimates caution against extrapolating evidence across cancer types. Cancer-specific evidence, endpoint definitions, and robustness analyses should therefore inform decisions to use late-stage incidence as an earlier trial end point.

The reduction from 26 retained publications to 18 unique trials and eight compatible comparisons shows why publication recovery is not equivalent to evidence expansion. Grouping publications by trial family reduced the risk of treating multiple reports from one trial as independent evidence. Independent source verification replaced the all-age Swedish Two-County mortality counts of 269/277 with age-compatible counts of 224/238, while the late-stage counts remained 389/432. Thus, the repeated agreement within one LLM configuration does not establish source accuracy.[29, 30]

A strength of this study was its end-to-end human verification of model-prioritized reports. Earlier LLM evaluations have concentrated mainly on title, abstract, or full-text classification against existing review labels.[10] This audit separately assessed publication-level relevance, trial identity, endpoint availability, and quantitative eligibility. Two additional reviewers independently verified all eight comparisons against the publications. The broader contribution is therefore to shift evaluation from agreement with inherited decisions to the validity of the evidence that ultimately enters a synthesis. Human verification becomes part of the analytic method rather than an administrative final check.[31, 32]

Several limitations affect interpretation. First, this was a sensitivity analysis, not formal surrogate validation;[33] cancer-specific strata were small, the 49 comparisons were equally weighted, and sampling error could substantially influence the correlations. The sparse colorectal comparison illustrates leverage, and no formal test compared the dependent original and updated correlations. Harmonized late-stage outcomes also included cancer-specific proxies (eg, tumor size, FIGO stage II or higher, Dukes C/D, and extensive-disease definitions), which may not be clinically equivalent across malignancies. The analysis did not reproduce the 63-data-point all-follow-up dataset or secondary definitions and follow-up pairings, and the descriptive I² summarized only five cancer-specific correlations. Significance concordance was secondary and limited by mixed derivation of the original and added classifications and by dichotomization at P < .05. Second, the audit was restricted to one review selected for reproducibility, one proprietary model and configuration, and a post hoc five-of-five threshold. Human adjudication did not cover 905 nonprioritized publications, 213 records without mapped PDFs, or the 58 reference-set publications, so true recall and false-negative frequency were unknown. PDF and supplement availability, English-language eligibility, possible model familiarity with the source review, and model updates may further limit generalizability and temporal reproducibility.[34] Future evaluations should include multiple reviews and models, post-cutoff evidence when available, and adjudicated samples of nonprioritized records.

The association between reductions in late-stage incidence and cancer-specific mortality was sensitive to evidence composition and varied by cancer type. LLM-assisted auditing may identify reports warranting reassessment, but human eligibility review, trial reconciliation, endpoint harmonization, and source verification determine whether those reports become usable evidence. These findings support cancer-specific, robustness-based evaluation of late-stage incidence as an earlier screening-trial end point.

## Data Availability

Data are available in a public, open-access repository. Derived audit data, the complete prompts, structured model outputs, analysis code, and computational environment information are available in the Open Science Framework at https://osf.io/jq3nw/overview.

## Data Availability

The data supporting the findings of this study are available at https://osf.io/jq3nw/overview.

## Acknowledgements.

OpenAI’s ChatGPT 5.6 was used to improve the writing of this manuscript and to assist with coding tasks. All AI-assisted text and code were reviewed, edited, and verified by the authors. The funders had no role in study design; data collection, analysis, or interpretation; manuscript preparation; or the decision to submit the manuscript for publication.

## Ethics approval

Ethics approval was not required because this methodological study analyzed published aggregate data and did not involve human participants, identifiable information, or individual participant data.

## Data Availability Statement

Data are available in a public, open-access repository. Derived audit data, the complete prompts, structured model outputs, analysis code, and computational environment information are available in the Open Science Framework at https://osf.io/jq3nw/overview. Source PDFs are not redistributed because access remains governed by their publishers. Supplementary Table S2 contains the prioritized-report inventory; Supplementary Table S3 reports independent human reassessment; Supplementary Table S4 reports endpoint auditing and trial-family reconciliation; Supplementary Table S5 reports independent source verification for the eight additions; and Supplementary Table S6 contains the complete 49-comparison dataset used to reproduce the results.

