## supplement for "LLM-assisted evidence audit of late-stage cancer incidence as a screening trial endpoint"

**Supplementary Material**

**Supplementary Table S1. Distribution of five-run LLM verdict patterns among the 996 publications with mapped full-text PDFs.**

| **Five-run verdict pattern** | **Publications, n** | **Inclusion-type decisions, n/5** | **Meets final five-of-five rule** |
| --- | --- | --- | --- |
| EXCLUDE, EXCLUDE, EXCLUDE, EXCLUDE, EXCLUDE | 855 | 0 | No |
| INCLUDE, INCLUDE, INCLUDE, INCLUDE, INCLUDE | 82 | 5 | Yes |
| EXCLUDE, EXCLUDE, EXCLUDE, EXCLUDE, Error | 21 | 0 | No |
| INCLUDE_DATA_IN_SUPPLEMENT, INCLUDE_DATA_IN_SUPPLEMENT, INCLUDE_DATA_IN_SUPPLEMENT, INCLUDE_DATA_IN_SUPPLEMENT, INCLUDE_DATA_IN_SUPPLEMENT | 7 | 5 | Yes |
| EXCLUDE, EXCLUDE, EXCLUDE, EXCLUDE, INCLUDE | 7 | 1 | No |
| Error, INCLUDE, INCLUDE, INCLUDE, INCLUDE | 6 | 4 | No |
| EXCLUDE, EXCLUDE, INCLUDE, INCLUDE, INCLUDE | 5 | 3 | No |
| EXCLUDE, INCLUDE, INCLUDE, INCLUDE, INCLUDE | 3 | 4 | No |
| EXCLUDE, EXCLUDE, EXCLUDE, EXCLUDE, INCLUDE_DATA_IN_SUPPLEMENT | 3 | 1 | No |
| EXCLUDE, EXCLUDE, EXCLUDE, Error, Error | 2 | 0 | No |
| EXCLUDE, EXCLUDE, EXCLUDE, INCLUDE, INCLUDE | 1 | 2 | No |
| INCLUDE, INCLUDE, INCLUDE, INCLUDE_DATA_IN_SUPPLEMENT, INCLUDE_DATA_IN_SUPPLEMENT | 1 | 5 | Yes |
| EXCLUDE, EXCLUDE, INCLUDE_DATA_IN_SUPPLEMENT, INCLUDE_DATA_IN_SUPPLEMENT, INCLUDE_DATA_IN_SUPPLEMENT | 1 | 3 | No |
| EXCLUDE, EXCLUDE, EXCLUDE, INCLUDE_DATA_IN_SUPPLEMENT, INCLUDE_DATA_IN_SUPPLEMENT | 1 | 2 | No |
| INCLUDE, INCLUDE, INCLUDE_DATA_IN_SUPPLEMENT, INCLUDE_DATA_IN_SUPPLEMENT, INCLUDE_DATA_IN_SUPPLEMENT | 1 | 5 | Yes |

*INCLUDE and INCLUDE_DATA_IN_SUPPLEMENT were counted as inclusion-type decisions; EXCLUDE and Error were not. Any Error therefore prevented a publication from meeting the five-of-five rule. The 15 patterns sum to 996 publications; the four retained patterns sum to 91.*

**Supplementary Table S2. Publication-level inventory of the 91 reports retained under the five-of-five LLM consensus rule.**

| **PMID** | **Year** | **Cancer type** | **Article title** | **Journal** | **Reference-set status** | **Five-run LLM verdict pattern** |
| --- | --- | --- | --- | --- | --- | --- |
| 3142562 | 1988 | Breast | Mammographic screening and mortality from breast cancer: the Malmö mammographic screening trial. | BMJ (Clinical research ed.) | In reference set | INCLUDE, INCLUDE, INCLUDE, INCLUDE, INCLUDE |
| 3411625 | 1988 | Breast | Analysis of breast cancer mortality and stage distribution by age for the Health Insurance Plan clinical trial. | Journal of the National Cancer Institute | Absent from reference set | INCLUDE, INCLUDE, INCLUDE, INCLUDE, INCLUDE |
| 1967717 | 1990 | Breast | Edinburgh trial of screening for breast cancer: mortality at seven years. | Lancet (London, England) | In reference set | INCLUDE, INCLUDE, INCLUDE, INCLUDE, INCLUDE |
| 1854979 | 1991 | Breast | Randomized study of mammography screening--preliminary report on mortality in the Stockholm trial. | Breast cancer research and treatment | Absent from reference set | INCLUDE, INCLUDE, INCLUDE, INCLUDE, INCLUDE |
| 2035504 | 1991 | Breast | Analysis of the temporal patterns of benefits in the Health Insurance Plan of Greater New York trial by stage and age. | American journal of epidemiology | Absent from reference set | INCLUDE, INCLUDE, INCLUDE, INCLUDE, INCLUDE |
| 8080744 | 1994 | Breast | The Edinburgh randomised trial of breast cancer screening: results after 10 years of follow-up. | British journal of cancer | In reference set | INCLUDE, INCLUDE, INCLUDE, INCLUDE, INCLUDE |
| 7497137 | 1995 | Breast | Effect of breast cancer screening after age 65. | Journal of medical screening | Absent from reference set | INCLUDE, INCLUDE, INCLUDE, INCLUDE, INCLUDE |
| 7579497 | 1995 | Breast | Nonattendance in the Stockholm mammography screening trial: relative mortality and reasons for nonattendance. | Breast cancer research and treatment | Absent from reference set | INCLUDE, INCLUDE, INCLUDE, INCLUDE, INCLUDE |
| 7736395 | 1995 | Breast | Efficacy of breast cancer screening by age. New results from the Swedish Two-County Trial. | Cancer | Absent from reference set | INCLUDE, INCLUDE, INCLUDE, INCLUDE, INCLUDE |
| 9386870 | 1997 | Breast | Followup after 11 years--update of mortality results in the Stockholm mammographic screening trial. | Breast cancer research and treatment | Absent from reference set | INCLUDE, INCLUDE, INCLUDE, INCLUDE, INCLUDE |
| 10995804 | 2000 | Breast | Canadian National Breast Screening Study-2: 13-year results of a randomized trial in women aged 50-59 years. | Journal of the National Cancer Institute | In reference set | INCLUDE, INCLUDE, INCLUDE, INCLUDE, INCLUDE |
| 12204013 | 2002 | Breast | The Canadian National Breast Screening Study-1: breast cancer mortality after 11 to 16 years of follow-up. A randomized screening trial of mammography in women age 40 to 49 years. | Annals of internal medicine | In reference set | INCLUDE, INCLUDE, INCLUDE, INCLUDE, INCLUDE |
| 26095467 | 2015 | Multiple / occult cancers | Screening for Occult Cancer in Unprovoked Venous Thromboembolism. | The New England journal of medicine | Absent from reference set | INCLUDE, INCLUDE, INCLUDE, INCLUDE_DATA_IN_SUPPLEMENT, INCLUDE_DATA_IN_SUPPLEMENT |
| 31221620 | 2019 | Breast | MRI versus mammography for breast cancer screening in women with familial risk (FaMRIsc): a multicentre, randomised, controlled trial. | The Lancet. Oncology | Absent from reference set | INCLUDE, INCLUDE, INCLUDE, INCLUDE, INCLUDE |
| 33141657 | 2020 | Breast | Annual mammographic screening to reduce breast cancer mortality in women from age 40 years: long-term follow-up of the UK Age RCT. | Health technology assessment (Winchester, England) | Absent from reference set | INCLUDE, INCLUDE, INCLUDE, INCLUDE, INCLUDE |
| 33627312 | 2021 | Breast | Effect of screening by clinical breast examination on breast cancer incidence and mortality after 20 years: prospective, cluster randomised controlled trial in Mumbai. | BMJ (Clinical research ed.) | In reference set | INCLUDE, INCLUDE, INCLUDE, INCLUDE, INCLUDE |
| 36321193 | 2023 | Breast | Effectiveness of triennial screening with clinical breast examination: 14-years follow-up outcomes of randomized clinical trial in Trivandrum, India. | Cancer | In reference set | INCLUDE, INCLUDE, INCLUDE, INCLUDE, INCLUDE |
| 19339719 | 2009 | Cervical | HPV screening for cervical cancer in rural India. | The New England journal of medicine | Absent from reference set | INCLUDE, INCLUDE, INCLUDE, INCLUDE, INCLUDE |
| 2762760 | 1989 | Colorectal | Repeated screening for colorectal cancer with fecal occult blood test. A prospective randomized study at Funen, Denmark. | Scandinavian journal of gastroenterology | In reference set | INCLUDE, INCLUDE, INCLUDE, INCLUDE, INCLUDE |
| 1290631 | 1992 | Prostate | Repeated screening for carcinoma of the prostate by digital rectal examination in a randomly selected population. | Acta oncologica (Stockholm, Sweden) | Absent from reference set | INCLUDE, INCLUDE, INCLUDE, INCLUDE, INCLUDE |
| 8340943 | 1993 | Colorectal | Screening for colorectal cancer with fecal occult blood testing and sigmoidoscopy. | Journal of the National Cancer Institute | Absent from reference set | INCLUDE, INCLUDE, INCLUDE, INCLUDE, INCLUDE |
| 8474513 | 1993 | Colorectal | Reducing mortality from colorectal cancer by screening for fecal occult blood. Minnesota Colon Cancer Control Study. | The New England journal of medicine | In reference set | INCLUDE, INCLUDE, INCLUDE, INCLUDE, INCLUDE |
| 8898422 | 1996 | Colorectal | Polypectomy of adenomas in the prevention of colorectal cancer: 10 years' follow-up of the Telemark Polyp Study I. A prospective, controlled population study. | Scandinavian journal of gastroenterology | Absent from reference set | INCLUDE, INCLUDE, INCLUDE, INCLUDE, INCLUDE |
| 8942774 | 1996 | Colorectal | Randomised study of screening for colorectal cancer with faecal-occult-blood test. | Lancet (London, England) | In reference set | INCLUDE, INCLUDE, INCLUDE, INCLUDE, INCLUDE |
| 8942775 | 1996 | Colorectal | Randomised controlled trial of faecal-occult-blood screening for colorectal cancer. | Lancet (London, England) | In reference set | INCLUDE, INCLUDE, INCLUDE, INCLUDE, INCLUDE |
| 10365903 | 1999 | Colorectal | Population-based surveillance by colonoscopy: effect on the incidence of colorectal cancer. Telemark Polyp Study I. | Scandinavian journal of gastroenterology | Absent from reference set | INCLUDE, INCLUDE, INCLUDE, INCLUDE, INCLUDE |
| 11772964 | 2002 | Colorectal | Protective effect of faecal occult blood test screening for colorectal cancer: worse prognosis for screening refusers. | Gut | In reference set | INCLUDE, INCLUDE, INCLUDE, INCLUDE, INCLUDE |
| 15188160 | 2004 | Colorectal | Reduction in colorectal cancer mortality by fecal occult blood screening in a French controlled study. | Gastroenterology | In reference set | INCLUDE, INCLUDE, INCLUDE, INCLUDE, INCLUDE |
| 18563785 | 2008 | Colorectal | Survival benefit in a randomized clinical trial of faecal occult blood screening for colorectal cancer. | The British journal of surgery | In reference set | INCLUDE, INCLUDE, INCLUDE, INCLUDE, INCLUDE |
| 19483252 | 2009 | Colorectal | Risk of colorectal cancer seven years after flexible sigmoidoscopy screening: randomised controlled trial. | BMJ (Clinical research ed.) | In reference set | INCLUDE, INCLUDE, INCLUDE, INCLUDE, INCLUDE |
| 21642681 | 2011 | Ovarian | Effect of screening on ovarian cancer mortality: the Prostate, Lung, Colorectal and Ovarian (PLCO) Cancer Screening Randomized Controlled Trial. | JAMA | In reference set | INCLUDE, INCLUDE, INCLUDE, INCLUDE, INCLUDE |
| 21852264 | 2011 | Colorectal | Once-only sigmoidoscopy in colorectal cancer screening: follow-up findings of the Italian Randomized Controlled Trial--SCORE. | Journal of the National Cancer Institute | In reference set | INCLUDE, INCLUDE, INCLUDE, INCLUDE, INCLUDE |
| 22031728 | 2011 | Lung | Screening by chest radiograph and lung cancer mortality: the Prostate, Lung, Colorectal, and Ovarian (PLCO) randomized trial. | JAMA | In reference set | INCLUDE, INCLUDE, INCLUDE, INCLUDE, INCLUDE |
| 22228146 | 2012 | Prostate | Prostate cancer screening in the randomized Prostate, Lung, Colorectal, and Ovarian Cancer Screening Trial: mortality results after 13 years of follow-up. | Journal of the National Cancer Institute | In reference set | INCLUDE, INCLUDE, INCLUDE, INCLUDE, INCLUDE |
| 22612596 | 2012 | Colorectal | Colorectal-cancer incidence and mortality with screening flexible sigmoidoscopy. | The New England journal of medicine | In reference set | INCLUDE, INCLUDE, INCLUDE, INCLUDE, INCLUDE |
| 23065684 | 2013 | Ovarian | Potential effect of the risk of ovarian cancer algorithm (ROCA) on the mortality outcome of the Prostate, Lung, Colorectal and Ovarian (PLCO) trial. | International journal of cancer | Absent from reference set | INCLUDE, INCLUDE, INCLUDE, INCLUDE, INCLUDE |
| 25117129 | 2014 | Colorectal | Effect of flexible sigmoidoscopy screening on colorectal cancer incidence and mortality: a randomized clinical trial. | JAMA | In reference set | INCLUDE, INCLUDE, INCLUDE, INCLUDE, INCLUDE |
| 28976536 | 2017 | Colorectal | Early detection versus primary prevention in the PLCO flexible sigmoidoscopy screening trial: Which has the greatest impact on mortality? | Cancer | Absent from reference set | INCLUDE_DATA_IN_SUPPLEMENT, INCLUDE_DATA_IN_SUPPLEMENT, INCLUDE_DATA_IN_SUPPLEMENT, INCLUDE_DATA_IN_SUPPLEMENT, INCLUDE_DATA_IN_SUPPLEMENT |
| 30288918 | 2019 | Prostate | Extended follow-up for prostate cancer incidence and mortality among participants in the Prostate, Lung, Colorectal and Ovarian randomized cancer screening trial. | BJU international | In reference set | INCLUDE, INCLUDE, INCLUDE, INCLUDE, INCLUDE |
| 30502933 | 2019 | Colorectal | Effect of flexible sigmoidoscopy screening on colorectal cancer incidence and mortality: long-term follow-up of the randomised US PLCO cancer screening trial. | The lancet. Gastroenterology & hepatology | In reference set | INCLUDE, INCLUDE, INCLUDE, INCLUDE, INCLUDE |
| 36214590 | 2022 | Colorectal | Effect of Colonoscopy Screening on Risks of Colorectal Cancer and Related Death. | The New England journal of medicine | In reference set | INCLUDE_DATA_IN_SUPPLEMENT, INCLUDE_DATA_IN_SUPPLEMENT, INCLUDE_DATA_IN_SUPPLEMENT, INCLUDE_DATA_IN_SUPPLEMENT, INCLUDE_DATA_IN_SUPPLEMENT |
| 14738659 | 2003 | Liver | Screening for liver cancer: results of a randomised controlled trial in Qidong, China. | Journal of medical screening | In reference set | INCLUDE, INCLUDE, INCLUDE, INCLUDE, INCLUDE |
| 15042359 | 2004 | Liver | Randomized controlled trial of screening for hepatocellular carcinoma. | Journal of cancer research and clinical oncology | In reference set | INCLUDE, INCLUDE, INCLUDE, INCLUDE, INCLUDE |
| 23750991 | 2013 | Liver | Surveillance for hepatocellular cancer with ultrasonography vs. computed tomography -- a randomised study. | Alimentary pharmacology & therapeutics | Absent from reference set | INCLUDE, INCLUDE, INCLUDE, INCLUDE, INCLUDE |
| 6734291 | 1984 | Lung | Screening for early lung cancer. Results of the Memorial Sloan-Kettering study in New York. | Chest | In reference set | INCLUDE, INCLUDE, INCLUDE, INCLUDE, INCLUDE |
| 2404878 | 1990 | Lung | Lack of benefit from semi-annual screening for cancer of the lung: follow-up report of a randomized controlled trial on a population of high-risk males in Czechoslovakia. | International journal of cancer | Absent from reference set | INCLUDE, INCLUDE, INCLUDE, INCLUDE, INCLUDE |
| 10944552 | 2000 | Lung | Lung cancer mortality in the Mayo Lung Project: impact of extended follow-up. | Journal of the National Cancer Institute | In reference set | INCLUDE, INCLUDE, INCLUDE, INCLUDE, INCLUDE |
| 11147611 | 2000 | Lung | The Mayo Lung Project: a perspective. | Cancer | Absent from reference set | INCLUDE, INCLUDE, INCLUDE, INCLUDE, INCLUDE |
| 11147613 | 2000 | Lung | Czech Study on Lung Cancer Screening: post-trial follow-up of lung cancer deaths up to year 15 since enrollment. | Cancer | Absent from reference set | INCLUDE, INCLUDE, INCLUDE, INCLUDE, INCLUDE |
| 19520905 | 2009 | Lung | A randomized study of lung cancer screening with spiral computed tomography: three-year results from the DANTE trial. | American journal of respiratory and critical care medicine | In reference set | INCLUDE, INCLUDE, INCLUDE, INCLUDE, INCLUDE |
| 19637354 | 2009 | Lung | Randomized controlled trials of the efficacy of lung cancer screening by sputum cytology revisited: a combined mortality analysis from the Johns Hopkins Lung Project and the Memorial Sloan-Kettering Lung Study. | Cancer | Absent from reference set | INCLUDE, INCLUDE, INCLUDE, INCLUDE, INCLUDE |
| 21714641 | 2011 | Lung | Reduced lung-cancer mortality with low-dose computed tomographic screening. | The New England journal of medicine | In reference set | INCLUDE, INCLUDE, INCLUDE, INCLUDE, INCLUDE |
| 22286927 | 2012 | Lung | CT screening for lung cancer brings forward early disease. The randomised Danish Lung Cancer Screening Trial: status after five annual screening rounds with low-dose CT. | Thorax | In reference set | INCLUDE, INCLUDE, INCLUDE, INCLUDE, INCLUDE |
| 25372087 | 2014 | Lung | Cost-effectiveness of CT screening in the National Lung Screening Trial. | The New England journal of medicine | Absent from reference set | INCLUDE_DATA_IN_SUPPLEMENT, INCLUDE_DATA_IN_SUPPLEMENT, INCLUDE_DATA_IN_SUPPLEMENT, INCLUDE_DATA_IN_SUPPLEMENT, INCLUDE_DATA_IN_SUPPLEMENT |
| 25760561 | 2015 | Lung | Long-Term Follow-up Results of the DANTE Trial, a Randomized Study of Lung Cancer Screening with Spiral Computed Tomography. | American journal of respiratory and critical care medicine | In reference set | INCLUDE, INCLUDE, INCLUDE, INCLUDE, INCLUDE |
| 26485620 | 2016 | Lung | Results of the Randomized Danish Lung Cancer Screening Trial with Focus on High-Risk Profiling. | American journal of respiratory and critical care medicine | In reference set | INCLUDE, INCLUDE, INCLUDE, INCLUDE, INCLUDE |
| 28377492 | 2017 | Lung | Mortality, survival and incidence rates in the ITALUNG randomised lung cancer screening trial. | Thorax | In reference set | INCLUDE, INCLUDE, INCLUDE, INCLUDE, INCLUDE |
| 30937431 | 2019 | Lung | Prolonged lung cancer screening reduced 10-year mortality in the MILD trial: new confirmation of lung cancer screening efficacy. | Annals of oncology : official journal of the European Society for Medical Oncology | In reference set | INCLUDE, INCLUDE, INCLUDE, INCLUDE, INCLUDE |
| 31260833 | 2019 | Lung | Lung Cancer Incidence and Mortality with Extended Follow-up in the National Lung Screening Trial. | Journal of thoracic oncology : official publication of the International Association for the Study of Lung Cancer | In reference set | INCLUDE, INCLUDE, INCLUDE, INCLUDE, INCLUDE |
| 31336289 | 2019 | Lung | Ten-year results of the Multicentric Italian Lung Detection trial demonstrate the safety and efficacy of biennial lung cancer screening. | European journal of cancer (Oxford, England : 1990) | Absent from reference set | INCLUDE, INCLUDE, INCLUDE_DATA_IN_SUPPLEMENT, INCLUDE_DATA_IN_SUPPLEMENT, INCLUDE_DATA_IN_SUPPLEMENT |
| 31537697 | 2019 | Lung | Sequential screening for lung cancer in a high-risk group: randomised controlled trial: LungSEARCH: a randomised controlled trial of Surveillance using sputum and imaging for the EARly detection of lung Cancer in a High-risk group. | The European respiratory journal | Absent from reference set | INCLUDE, INCLUDE, INCLUDE, INCLUDE, INCLUDE |
| 31162856 | 2020 | Lung | Lung cancer mortality reduction by LDCT screening-Results from the randomized German LUSI trial. | International journal of cancer | In reference set | INCLUDE_DATA_IN_SUPPLEMENT, INCLUDE_DATA_IN_SUPPLEMENT, INCLUDE_DATA_IN_SUPPLEMENT, INCLUDE_DATA_IN_SUPPLEMENT, INCLUDE_DATA_IN_SUPPLEMENT |
| 31995683 | 2020 | Lung | Reduced Lung-Cancer Mortality with Volume CT Screening in a Randomized Trial. | The New England journal of medicine | In reference set | INCLUDE, INCLUDE, INCLUDE, INCLUDE, INCLUDE |
| 32732334 | 2021 | Lung | Earlier diagnosis of lung cancer in a randomised trial of an autoantibody blood test followed by imaging. | The European respiratory journal | In reference set | INCLUDE, INCLUDE, INCLUDE, INCLUDE, INCLUDE |
| 31373615 | 2019 | Nasopharyngeal | Incidence and mortality of nasopharyngeal carcinoma: interim analysis of a cluster randomized controlled screening trial (PRO-NPC-001) in southern China. | Annals of oncology : official journal of the European Society for Medical Oncology | In reference set | INCLUDE, INCLUDE, INCLUDE, INCLUDE, INCLUDE |
| 12798401 | 2003 | Oral | Interim results from a cluster randomized controlled oral cancer screening trial in Kerala, India. | Oral oncology | In reference set | INCLUDE, INCLUDE, INCLUDE, INCLUDE, INCLUDE |
| 15936419 | 2005 | Oral | Effect of screening on oral cancer mortality in Kerala, India: a cluster-randomised controlled trial. | Lancet (London, England) | In reference set | INCLUDE, INCLUDE, INCLUDE, INCLUDE, INCLUDE |
| 10217079 | 1999 | Ovarian | Screening for ovarian cancer: a pilot randomised controlled trial. | Lancet (London, England) | In reference set | INCLUDE, INCLUDE, INCLUDE, INCLUDE, INCLUDE |
| 26707054 | 2016 | Ovarian | Ovarian cancer screening and mortality in the UK Collaborative Trial of Ovarian Cancer Screening (UKCTOCS): a randomised controlled trial. | Lancet (London, England) | In reference set | INCLUDE_DATA_IN_SUPPLEMENT, INCLUDE_DATA_IN_SUPPLEMENT, INCLUDE_DATA_IN_SUPPLEMENT, INCLUDE_DATA_IN_SUPPLEMENT, INCLUDE_DATA_IN_SUPPLEMENT |
| 33991479 | 2021 | Ovarian | Ovarian cancer population screening and mortality after long-term follow-up in the UK Collaborative Trial of Ovarian Cancer Screening (UKCTOCS): a randomised controlled trial. | Lancet (London, England) | In reference set | INCLUDE, INCLUDE, INCLUDE, INCLUDE, INCLUDE |
| 37657461 | 2023 | Ovarian | Tumour stage, treatment, and survival of women with high-grade serous tubo-ovarian cancer in UKCTOCS: an exploratory analysis of a randomised controlled trial. | The Lancet. Oncology | Absent from reference set | INCLUDE, INCLUDE, INCLUDE, INCLUDE, INCLUDE |
| 15548438 | 2004 | Prostate | Clinical consequences of screening for prostate cancer: 15 years follow-up of a randomised controlled trial in Sweden. | European urology | In reference set | INCLUDE, INCLUDE, INCLUDE, INCLUDE, INCLUDE |
| 19297565 | 2009 | Prostate | Mortality results from a randomized prostate-cancer screening trial. | The New England journal of medicine | In reference set | INCLUDE, INCLUDE, INCLUDE, INCLUDE, INCLUDE |
| 19297566 | 2009 | Prostate | Screening and prostate-cancer mortality in a randomized European study. | The New England journal of medicine | In reference set | INCLUDE_DATA_IN_SUPPLEMENT, INCLUDE_DATA_IN_SUPPLEMENT, INCLUDE_DATA_IN_SUPPLEMENT, INCLUDE_DATA_IN_SUPPLEMENT, INCLUDE_DATA_IN_SUPPLEMENT |
| 20598634 | 2010 | Prostate | Mortality results from the Göteborg randomised population-based prostate-cancer screening trial. | The Lancet. Oncology | In reference set | INCLUDE, INCLUDE, INCLUDE, INCLUDE, INCLUDE |
| 21454449 | 2011 | Prostate | Randomised prostate cancer screening trial: 20 year follow-up. | BMJ (Clinical research ed.) | In reference set | INCLUDE, INCLUDE, INCLUDE, INCLUDE, INCLUDE |
| 22417251 | 2012 | Prostate | Prostate-cancer mortality at 11 years of follow-up. | The New England journal of medicine | In reference set | INCLUDE, INCLUDE, INCLUDE, INCLUDE, INCLUDE |
| 23479454 | 2013 | Prostate | Prostate cancer mortality in the Finnish randomized screening trial. | Journal of the National Cancer Institute | In reference set | INCLUDE, INCLUDE, INCLUDE, INCLUDE, INCLUDE |
| 23759326 | 2013 | Prostate | Screening for prostate cancer: results of the Rotterdam section of the European randomized study of screening for prostate cancer. | European urology | In reference set | INCLUDE, INCLUDE, INCLUDE, INCLUDE, INCLUDE |
| 25108889 | 2014 | Prostate | Screening and prostate cancer mortality: results of the European Randomised Study of Screening for Prostate Cancer (ERSPC) at 13 years of follow-up. | Lancet (London, England) | In reference set | INCLUDE, INCLUDE, INCLUDE, INCLUDE, INCLUDE |
| 25556937 | 2015 | Prostate | Opportunistic testing versus organized prostate-specific antigen screening: outcome after 18 years in the Göteborg randomized population-based prostate cancer screening trial. | European urology | Absent from reference set | INCLUDE, INCLUDE, INCLUDE, INCLUDE, INCLUDE |
| 25698407 | 2015 | Prostate | Do Treatment Differences between Arms Affect the Main Outcome of ERSPC Rotterdam? | The Journal of urology | Absent from reference set | INCLUDE, INCLUDE, INCLUDE, INCLUDE, INCLUDE |
| 25777669 | 2015 | Prostate | Update of the results of the Spanish branch of the European Randomized Study on Screening for Prostate Cancer (ERSPC). | Actas urologicas espanolas | Absent from reference set | INCLUDE, INCLUDE, INCLUDE, INCLUDE, INCLUDE |
| 29224236 | 2018 | Prostate | Prognostic factors of prostate cancer mortality in a Finnish randomized screening trial. | International journal of urology : official journal of the Japanese Urological Association | Absent from reference set | INCLUDE, INCLUDE, INCLUDE, INCLUDE, INCLUDE |
| 29254399 | 2018 | Prostate | Eighteen-year follow-up of the Göteborg Randomized Population-based Prostate Cancer Screening Trial: effect of sociodemographic variables on participation, prostate cancer incidence and mortality. | Scandinavian journal of urology | In reference set | INCLUDE, INCLUDE, INCLUDE, INCLUDE, INCLUDE |
| 29509864 | 2018 | Prostate | Effect of a Low-Intensity PSA-Based Screening Intervention on Prostate Cancer Mortality: The CAP Randomized Clinical Trial. | JAMA | In reference set | INCLUDE, INCLUDE, INCLUDE, INCLUDE, INCLUDE |
| 30653262 | 2019 | Prostate | Bias-corrected estimates of effects of PSA screening decisions on the risk of prostate cancer diagnosis and death: Analysis of the Finnish randomized study of screening for prostate cancer. | International journal of cancer | Absent from reference set | INCLUDE, INCLUDE, INCLUDE, INCLUDE, INCLUDE |
| 30824296 | 2019 | Prostate | A 16-yr Follow-up of the European Randomized study of Screening for Prostate Cancer. | European urology | In reference set | INCLUDE, INCLUDE, INCLUDE, INCLUDE, INCLUDE |
| 34817559 | 2022 | Prostate | Outcomes of Screening for Prostate Cancer Among Men Who Use Statins. | JAMA oncology | Absent from reference set | INCLUDE_DATA_IN_SUPPLEMENT, INCLUDE_DATA_IN_SUPPLEMENT, INCLUDE_DATA_IN_SUPPLEMENT, INCLUDE_DATA_IN_SUPPLEMENT, INCLUDE_DATA_IN_SUPPLEMENT |
| 35422134 | 2022 | Prostate | Results from 22 years of Followup in the Göteborg Randomized Population-Based Prostate Cancer Screening Trial. | The Journal of urology | In reference set | INCLUDE, INCLUDE, INCLUDE, INCLUDE, INCLUDE |
| 37029074 | 2023 | Prostate | A Detailed Evaluation of the Effect of Prostate-specific Antigen-based Screening on Morbidity and Mortality of Prostate Cancer: 21-year Follow-up Results of the Rotterdam Section of the European Randomised Study of Screening for Prostate Cancer. | European urology | Absent from reference set | INCLUDE, INCLUDE, INCLUDE, INCLUDE, INCLUDE |

**Supplementary Table S3. Independent human reassessment and final audit disposition of the 33 LLM-retained publications absent from the reference set.**

| **PMID** | **Reviewer 1 decision** | **Reviewer 1 rationale** | **Reviewer 2 decision** | **Reviewer 2 rationale** | **Initial reviewer agreement** | **Resolution pathway** | **Final human disposition** | **Final rationale** |
| --- | --- | --- | --- | --- | --- | --- | --- | --- |
| 3411625 | Retain for reconciliation | — | Retain for reconciliation | — | Yes | No adjudication required | Retain for trial reconciliation | Eligible on human re-review; publication was judged to meet the operational screening-review criteria. |
| 1854979 | Retain for reconciliation | — | Retain for reconciliation | — | Yes | No adjudication required | Retain for trial reconciliation | Eligible on human re-review; publication was judged to meet the operational screening-review criteria. |
| 2035504 | Retain for reconciliation | — | Retain for reconciliation | — | Yes | No adjudication required | Retain for trial reconciliation | Eligible on human re-review; publication was judged to meet the operational screening-review criteria. |
| 7497137 | Retain for reconciliation | — | Retain for reconciliation | — | Yes | No adjudication required | Retain for trial reconciliation | Eligible on human re-review; publication was judged to meet the operational screening-review criteria. |
| 7579497 | Retain for reconciliation | WHO histopathological criteria (stages I-IV). | Retain for reconciliation | — | Yes | No adjudication required | Retain for trial reconciliation | Eligible on human re-review; publication was judged to meet the operational screening-review criteria. |
| 7736395 | Retain for reconciliation | Tumor size? | Retain for reconciliation | — | Yes | No adjudication required | Retain for trial reconciliation | Eligible on human re-review; late-stage disease judged mappable from tumor size. |
| 9386870 | Retain for reconciliation | — | Retain for reconciliation | — | Yes | No adjudication required | Retain for trial reconciliation | Eligible on human re-review; publication was judged to meet the operational screening-review criteria. |
| 26095467 | Retain for reconciliation | TNM classification system | Retain for reconciliation | — | Yes | No adjudication required | Retain for trial reconciliation | Eligible on human re-review; late-stage disease judged mappable from TNM classification. |
| 31221620 | Retain for reconciliation | — | Retain for reconciliation | — | Yes | No adjudication required | Retain for trial reconciliation | Eligible on human re-review; publication was judged to meet the operational screening-review criteria. |
| 33141657 | Retain for reconciliation | — | Retain for reconciliation | — | Yes | No adjudication required | Retain for trial reconciliation | Eligible on human re-review; publication was judged to meet the operational screening-review criteria. |
| 19339719 | Retain for reconciliation | — | Retain for reconciliation | — | Yes | No adjudication required | Retain for trial reconciliation | Eligible on human re-review; publication was judged to meet the operational screening-review criteria. |
| 1290631 | Retain for reconciliation | T0-T2, N0, M0,T3 and/or metastatic disease | Retain for reconciliation | — | Yes | No adjudication required | Retain for trial reconciliation | Eligible on human re-review; publication was judged to meet the operational screening-review criteria. |
| 8340943 | Retain for reconciliation | Dukes stage | Retain for reconciliation | — | Yes | Final design recheck | Exclude | Ineligible after final eligibility recheck; allocation by date rather than randomization did not satisfy the randomized controlled trial criterion, superseding the original endpoint-oriented Yes decisions. |
| 8898422 | Retain for reconciliation | Dukes stage | Retain for reconciliation | — | Yes | No adjudication required | Retain for trial reconciliation | Eligible on human re-review; late-stage disease judged mappable from Dukes classification. |
| 10365903 | Retain for reconciliation | Dukes stage | Retain for reconciliation | — | Yes | No adjudication required | Retain for trial reconciliation | Eligible on human re-review; late-stage disease judged mappable from Dukes classification. |
| 23065684 | Retain for reconciliation | TNM classification system | Retain for reconciliation | — | Yes | No adjudication required | Retain for trial reconciliation | Eligible on human re-review; late-stage disease judged mappable from TNM classification. |
| 28976536 | Retain for reconciliation | — | Retain for reconciliation | — | Yes | No adjudication required | Retain for trial reconciliation | Eligible on human re-review; publication was judged to meet the operational screening-review criteria. |
| 23750991 | Retain for reconciliation | — | Retain for reconciliation | — | Yes | No adjudication required | Retain for trial reconciliation | Eligible on human re-review; publication was judged to meet the operational screening-review criteria. |
| 2404878 | Retain for reconciliation | — | Retain for reconciliation | — | Yes | No adjudication required | Retain for trial reconciliation | Eligible on human re-review; publication was judged to meet the operational screening-review criteria. |
| 11147611 | Exclude | No classifiable late-stage endpoint | Exclude | — | Yes | No adjudication required | Exclude | Ineligible on human re-review; late-stage disease could not be categorized or extracted under the review rules. |
| 11147613 | Exclude | No classifiable late-stage endpoint | Retain for reconciliation | — | No | Adjudicated after reviewer disagreement | Retain for trial reconciliation | Eligible after reviewer disagreement; final adjudication accepted the stage mapping despite an initial categorization concern. |
| 19637354 | Retain for reconciliation | TNM classification system | Retain for reconciliation | — | Yes | No adjudication required | Retain for trial reconciliation | Eligible on human re-review; late-stage disease judged mappable from TNM classification. |
| 25372087 | Exclude | No classifiable late-stage endpoint | Retain for reconciliation | — | No | Adjudicated after reviewer disagreement | Exclude | Ineligible on human re-review; late-stage disease could not be categorized or extracted under the review rules. |
| 31336289 | Exclude | No classifiable late-stage endpoint; stage I–II and stage III–IV were stratified by intensity | Retain for reconciliation | — | No | Adjudicated after reviewer disagreement | Exclude | Ineligible on human re-review; late-stage disease could not be categorized or extracted under the review rules. |
| 31537697 | Retain for reconciliation | — | Retain for reconciliation | — | Yes | No adjudication required | Retain for trial reconciliation | Eligible on human re-review; publication was judged to meet the operational screening-review criteria. |
| 37657461 | Retain for reconciliation | — | Retain for reconciliation | — | Yes | No adjudication required | Retain for trial reconciliation | Eligible on human re-review; publication was judged to meet the operational screening-review criteria. |
| 25556937 | Exclude | No classifiable late-stage endpoint; stage I–II and stage III–IV were not separately categorized | Exclude | No stage classification reported | Yes | No adjudication required | Exclude | Ineligible on human re-review; late-stage disease could not be categorized or extracted under the review rules. |
| 25698407 | Retain for reconciliation | D'Amico criteria | Retain for reconciliation | — | Yes | No adjudication required | Retain for trial reconciliation | Eligible on human re-review; publication was judged to meet the operational screening-review criteria. |
| 25777669 | Exclude | Spanish-language report | Exclude | Spanish-language report | Yes | No adjudication required | Exclude | Ineligible on human re-review; excluded on language grounds. |
| 29224236 | Retain for reconciliation | TNM classification system | Retain for reconciliation | — | Yes | No adjudication required | Retain for trial reconciliation | Eligible on human re-review; late-stage disease judged mappable from TNM classification. |
| 30653262 | Retain for reconciliation | TNM classification | Retain for reconciliation | — | Yes | No adjudication required | Retain for trial reconciliation | Eligible on human re-review; late-stage disease judged mappable from TNM classification. |
| 34817559 | Retain for reconciliation | Gleason score, TNM | Retain for reconciliation | — | Yes | No adjudication required | Retain for trial reconciliation | Eligible on human re-review; late-stage disease judged mappable from TNM classification. |
| 37029074 | Exclude | No classifiable late-stage endpoint; stage I–II and stage III–IV were not separately categorized | Exclude | No stage classification reported | Yes | No adjudication required | Exclude | Ineligible on human re-review; late-stage disease could not be categorized or extracted under the review rules. |

*Initial reviewer decisions agreed for 30 reports and differed for three. After resolution, 26 reports were retained for unique-trial reconciliation and seven were excluded. PMID 8340943 underwent a final design recheck and was excluded because allocation by date did not satisfy the randomized-trial criterion, despite concordant initial reviewer decisions to retain.*

**Supplementary Table S4. Publication-level endpoint audit and unique-trial reconciliation for the 26 human-retained reports representing 18 unique trials.**

**Panel A. Publication-level endpoint extraction for 26 retained reports (one row per publication).**

| **Unique trial ID** | **PMID** | **Cancer type** | **Cancer-specific deaths I/C** | **Late-stage cases I/C** | **Number of runs returning the same four-count endpoint tuple** | **Source-check outcome** | **Endpoint usability** | **Issue code(s)** | **Source location(s)** |
| --- | --- | --- | --- | --- | --- | --- | --- | --- | --- |
| T01 | 11147613 | Lung | 247 / 216 | 53 / 46 | 5/5 | Accepted from unanimous 5/5 extraction | Supporting or later report | — | Mortality: Page 4, Results section 'Mortality from Lung Cancer' Late stage: Page 3, Results section 'Comparison of Staging, Resectability, and Survival of Cases of the Intervention versus Control Groups' |
| T01 | 2404878 | Lung | 85 / 67 | 53 / 46 | 5/5 | Accepted from unanimous 5/5 extraction | Selected analytic input | — | Mortality: Page 5, Results Section (also 64 vs 47 in Table I, Page 3) Late stage: Table II, Page 4 |
| T02 | 25698407 | Prostate | 151 / 188 | 720 / 694 | 5/5 | Accepted from unanimous 5/5 extraction | Not added—already represented | — | Mortality: Table 2, Page 5 Late stage: Table 2, Page 5 |
| T03 | 31221620 | Breast | 0 / 0 | 3 / 2 | 5/5 | Manual PDF verification completed | Not added—not poolable | M | Mortality: Outcomes section, page 5; Results section, page 9 Late stage: Table 2, page 6; T2 or higher invasive cancers |
| T04 | 29224236 | Prostate | 278 / 456 | 889 / 1675 | 5/5 | Accepted from unanimous 5/5 extraction | Not added—already represented | — | Mortality: Table 1, Page 3 Late stage: Table 1, Page 3 |
| T04 | 30653262 | Prostate | 77 / 152 | 167 / 519 | 2/5 | Manual PDF verification completed | Not added—already represented | F, M, L, R | Mortality: Table 3, page 6 Late stage: Table 2, page 5; corrected arms using progressed + metastatic disease |
| T04 | 34817559 | Prostate | 287 / 497 | Not Found / Not Found | 3/5 | Run disagreement; manual verification required | Not added—already represented | — | Mortality: Table 1, Page 4 (calculated by summing PCa mortality for statin users and nonusers in each arm) Late stage: eTable 1 and eTable 2 in the Supplement, Page 3 and 5 |
| T05 | 2035504 | Breast | 153 / 196 | 42 / 65 | 5/5 | Accepted from unanimous 5/5 extraction | Not added—not poolable | D, M | Mortality: Table 1 and Table 2 (18+ years columns summed), Page 4 Late stage: Table 1 and Table 2 (parenthetical values C3/C4 and S3/S4 summed), Page 4 |
| T05 | 3411625 | Breast | 153 / 196 | 42 / 65 | 5/5 | Manual PDF verification completed | Not added—not poolable | D, M | Mortality: Tables 1 and 2, page 3; study deaths 61+92 and control deaths 77+119 Late stage: Tables 1 and 2, page 3; study stage III-IV cases (12+8)+(12+10) and control stage III-IV cases (15+8)+(28+14) |
| T06 | 19637354 | Lung | 259 / 294 | 191 / 193 | 5/5 | Accepted from unanimous 5/5 extraction | Not added—already represented | — | Mortality: Table 5, Page 20 Late stage: Table 3, Page 16 |
| T07 | 31537697 | Lung | 16 / 21 | 19 / 17 | 5/5 | Accepted from unanimous 5/5 extraction | Selected analytic input | — | Mortality: Page 8, section 'Other cancers, mortality and smoking status' Late stage: Table 3 (Page 6) and Table 4 (Page 7) |
| T08 | 28976536 | Colorectal | 253 / 351 | / | 3/5 | Manual PDF verification completed | Not added—already represented | L, S, R | Mortality: Results section, page 4 Late stage: Results section, page 5; exact counts not present in the available PDF and the cited supplemental table is missing |
| T09 | 23065684 | Ovarian | 132 / 119 | 182 / 166 | 5/5 | Accepted from unanimous 5/5 extraction | Not added—already represented | — | Mortality: Table 1, Page 5 Late stage: Table 1, Page 5 |
| T10 | 23750991 | Liver | 5 / 7 | 4 / 3 | 4/5 | Manual PDF verification completed | Selected analytic input | A | Mortality: Table 3, page 4 Late stage: Table 3, page 4; summed TNM stage III, IIIA, and IVB |
| T11 | 19339719 | Cervical | 34 / 64 | 39 / 82 | 5/5 | Accepted from unanimous 5/5 extraction | Selected analytic input | — | Mortality: Table 4, Page 8 Late stage: Table 4, Page 8 and Abstract, Page 1 |
| T12 | 9386870 | Breast | 66 / 45 | 28 / 12 | 5/5 | Accepted from unanimous 5/5 extraction | Supporting or later report | — | Mortality: Table 2, Page 3 Late stage: Table 1, Page 2 |
| T12 | 1854979 | Breast | 39 / 30 | 34 / 39 | 3/5 | Manual PDF verification completed | Supporting or duplicate report | L | Mortality: Table 5, page 4 Late stage: Table 4, page 4 |
| T12 | 7579497 | Breast | 43 / 42 | 17 / 10 | 5/5 | Accepted from unanimous 5/5 extraction | Selected analytic input | — | Mortality: Table 2, Page 4 (calculated by summing No. breast cancer deaths for 'A+B' [23] and 'Invited screening' [20]) Late stage: Table 2, Page 4 (calculated by summing Stage III and Stage IV cases for groups A, B, and 'Invited screening') |
| T13 | 1290631 | Prostate | 2 / 7 | 6 / 32 | 5/5 | Accepted from unanimous 5/5 extraction | Not added—already represented | — | Mortality: Page 5, Results Section (Text) Late stage: Table 5, Page 5 |
| T14 | 7736395 | Breast | 269 / 277 | 389 / 432 | 5/5 | Corrected during duplicate reviewer audit | Supporting or later report | — | Mortality: Page 3 (Results Section) Late stage: Page 6, Table 4 |
| T14 | 7497137 | Breast | 224 / 238 | 389 / 432 | 3/5 | Manual PDF verification completed | Selected analytic input | F | Mortality: Table 1, page 2; summed age groups 50-64 and 65-74 to align with the stage table Late stage: Table 2, page 3; summed tumour-size categories >=20 mm across age groups 50-64 and 65-74 |
| T15 | 10365903 | Colorectal | 1 / 3 | 1 / 5 | 5/5 | Accepted from unanimous 5/5 extraction | Supporting or later report | — | Mortality: Table III and Table IV, Pages 5-6 Late stage: Table III (interpreted using Dukes mapping), Page 5 |
| T15 | 8898422 | Colorectal | 1 / 1 | 1 / 2 | 5/5 | Accepted from unanimous 5/5 extraction | Selected analytic input | — | Mortality: Table IV, Page 4 Late stage: Text (Colorectal cancer section) and Table V, Page 4 |
| T16 | 33141657 | Breast | 209 / 474 | 253 / 562 | 4/5 | Manual PDF verification completed | Selected analytic input | F | Mortality: Table 3, page 30 Late stage: Table 9, page 36; summed intervention screen-detected and symptomatic cancers with tumour size >20 mm |
| T17 | 37657461 | Ovarian | 205 / 446 | 195 / 446 | 3/5 | Manual PDF verification completed | Not added—already represented | D, L | Mortality: Abstract and Results text, page 6 Late stage: Table 1, page 4; advanced stage by intention to screen |
| T18 | 26095467 | Multiple / occult cancers | 4 / 6 | 16 / 13 | 1/5 | Run disagreement; manual verification required | Not added—not poolable | — | Mortality: Page 5, Results Section (Clinical Outcomes) Late stage: Page 3 (Outcome Assessment) and Page 5, Results Section (Clinical Outcomes) |

**Panel B. Unique-trial reconciliation for 18 unique trials (one row per unique trial).**

| **Unique trial ID** | **Unique trial** | **Cancer type** | **Human-retained PMID(s)** | **Relationship to original 41-comparison dataset** | **Selected analysis source (PMID)** | **Final analytic disposition** | **Selection rationale** |
| --- | --- | --- | --- | --- | --- | --- | --- |
| T01 | Czech Lung Screening Trial | Lung | 11147613, 2404878 | New audit-recovered trial comparison | 2404878 | Added | Earliest report with compatible randomized-arm mortality and stage data: deaths 85/67 and stage III cases 53/46. The later mortality follow-up in PMID 11147613 was not paired with a compatible stage-assessment window. |
| T02 | ERSPC Rotterdam treatment-difference analysis | Prostate | 25698407 | Overlaps an original comparison | — | Already represented | Same ERSPC Rotterdam trial as Feng-included PMID 23759326; do not add as an independent unique-trial row. |
| T03 | FaMRIsc Trial | Breast | 31221620 | Additional trial; not poolable | — | Not poolable | Single confirmed additional publication in this trial; screening-eligible and descriptively extractable, but not suitable for the mortality-reduction sensitivity update because randomized-arm breast-cancer mortality was not yet mature. |
| T04 | Finnish Randomized Study of Prostate Cancer Screening | Prostate | 29224236, 30653262, 34817559 | Overlaps an original comparison | — | Already represented | Same Finnish prostate-screening trial as Feng-included PMID 23479454. PMIDs 29224236, 30653262, and 34817559 are prognostic, bias-corrected, or subgroup/secondary analyses and are not independent unique-trial additions. |
| T05 | HIP Breast Screening Trial | Breast | 3411625, 2035504 | Additional trial; not poolable | — | Not poolable | Same HIP Breast Screening Trial as PMID 2035504; descriptively useful, but not suitable for denominator-based sensitivity pooling because the reported mortality counts are restricted to women diagnosed with breast cancer within 6 years of trial entry. |
| T06 | Johns Hopkins Lung Project / Memorial Sloan-Kettering Lung Study combined sputum-cytology analysis | Lung | 19637354 | Partial overlap with an original comparison | — | Already represented / not independently addable | PMID 19637354 combines the Johns Hopkins Lung Project with the Memorial Sloan-Kettering Lung Study, the latter represented in Feng et al. by PMID 6734291. It was not added as an independent comparison because separable compatible endpoint data were unavailable. |
| T07 | LungSEARCH Trial | Lung | 31537697 | New audit-recovered trial comparison | 31537697 | Added | Only retained report for this trial; compatible randomized-arm mortality and stage III–IV or extensive-disease counts were available. |
| T08 | PLCO Flexible Sigmoidoscopy Trial | Colorectal | 28976536 | Overlaps an original comparison | — | Already represented | Same PLCO Flexible Sigmoidoscopy Trial as Feng-included PMIDs 22612596 and 30502933. PMID 28976536 is a secondary mortality analysis, not an independent unique-trial addition; exact late-stage counts were unavailable in the current PDF. |
| T09 | PLCO ROCA ovarian-screening analysis | Ovarian | 23065684 | Overlaps an original comparison | — | Already represented | Same PLCO ovarian trial as Feng-included PMID 21642681; the ROCA analysis is not an independent unique-trial row. |
| T10 | Randomized Hepatocellular Carcinoma Surveillance Trial | Liver | 23750991 | New audit-recovered trial comparison | 23750991 | Added | Only retained report for this trial; source review confirmed ultrasonography as the intervention and computed tomography as the comparator. |
| T11 | Rural India HPV Screening Trial | Cervical | 19339719 | New audit-recovered trial comparison | 19339719 | Added | Only retained report for this trial; the HPV-screening versus standard-care comparison reported compatible mortality and FIGO stage II-or-higher data. |
| T12 | Stockholm Mammography Trial | Breast | 1854979, 7579497, 9386870 | New audit-recovered trial comparison | 7579497 | Added | Earliest compatible invited-versus-control report. The full intervention arm was reconstructed as screened women plus nonattenders; PMID 9386870 provided a later follow-up and was not selected. |
| T13 | Swedish / Norrköping prostate DRE screening trial | Prostate | 1290631 | Overlaps an original comparison | — | Already represented | Same Norrköping prostate screening trial as Feng-included PMIDs 15548438 and 21454449; do not add as an independent unique-trial row. |
| T14 | Swedish Two-County Trial | Breast | 7497137, 7736395 | New audit-recovered trial comparison | 7497137 | Added | Earliest compatible data for the same population aged 50–74 years: deaths 224/238 paired with tumours measuring at least 20 mm, 389/432. The later report PMID 7736395 was not selected. |
| T15 | Telemark Polyp Study I | Colorectal | 8898422, 10365903 | New audit-recovered trial comparison | 8898422 | Added | Earliest report with compatible randomized-arm colorectal-cancer mortality and Dukes C/D data. The later report PMID 10365903 was not selected. |
| T16 | UK Age RCT | Breast | 33141657 | New audit-recovered trial comparison | 33141657 | Added | Only retained report for this trial; compatible mortality and invasive-tumour-size data were available, with tumour diameter greater than 20 mm used as the prespecified breast-cancer proxy. |
| T17 | UKCTOCS | Ovarian | 37657461 | Overlaps an original comparison | — | Already represented | Same UKCTOCS trial as Feng-included PMIDs 26707054 and 33991479. PMID 37657461 is a histotype-restricted high-grade-serous tubo-ovarian subgroup analysis and is not an independent unique-trial addition. |
| T18 | Unprovoked VTE occult-cancer screening trial | Multiple / occult cancers | 26095467 | Additional trial; not poolable | — | Not poolable | Human-confirmed as audit-relevant, but not a site-specific cancer-screening trial input and not directly poolable in the Feng-style cancer-screening sensitivity update. |

*I/C denotes intervention/control. Endpoint counts in Panel A are audit extractions and are not automatically analytic inputs. Number of runs returning the same four-count endpoint tuple describes tuple agreement among the model runs before manual or duplicate source review and does not supersede a later source-based correction. Issue codes are A, arm reversal; F, wrong follow-up or analysis frame; D, subgroup denominator or restricted analysis population; M, mortality endpoint mismatch; L, late-stage mapping ambiguity; S, supplement-dependent extraction; and R, missing raw randomized-arm counts. A dash means that no prespecified issue code was assigned.*

**Supplementary Table S5. Source-verification and analytic selection status of the eight earliest-compatible trial-level comparisons included in the sensitivity analysis.**

| **Trial** | **Selected PMID** | **Intervention/control contrast** | **Final analytic endpoint tuple** | **Stage III–IV definition or cancer-specific proxy** | **Two-reviewer source check** | **Analytic selection or correction** | **Final analytic status** | **Source location(s)** |
| --- | --- | --- | --- | --- | --- | --- | --- | --- |
| Stockholm Mammography Trial | 7579497 | Invitation to mammographic screening, including screened women and nonattenders / noninvited control | Participants 40340 / 19943; cancer deaths 43 / 42; stage III–IV cases 17 / 10 | WHO stage III–IV | Source values confirmed by two reviewers | Author-selected the earliest-compatible tuple; the later tuple was 40318/19943; 66/45; 28/12 | Included using the author-selected earliest-compatible tuple | Table 2, PDF p 4 (journal p 270); intervention total reconstructed as screened women plus nonattenders |
| Swedish Two-County Trial | 7497137 | Invitation to mammographic screening / control, women aged 50–74 years | Participants 57236 / 40381; cancer deaths 224 / 238; stage III–IV cases 389 / 432 | Tumour diameter ≥20 mm (breast proxy) | Two-reviewer source check identified a mortality-count correction | Reviewer-identified correction: all-age deaths 269/277 were replaced with age-compatible deaths 224/238; stage counts remained 389/432 | Included after the two-reviewer correction | Table 1, PDF p 2: denominators and deaths; Table 2, PDF p 3: tumour size |
| UK Age RCT | 33141657 | Annual mammographic screening / usual care | Participants 53883 / 106953; cancer deaths 209 / 474; stage III–IV cases 253 / 562 | Invasive tumour diameter >20 mm (breast proxy) | Source values confirmed by two reviewers | None | Confirmed without correction; included | Abstract: denominators; Table 3, PDF p 30: mortality; Table 9, PDF p 36: tumour size >20 mm |
| Rural India HPV Screening Trial | 19339719 | Single-round HPV screening / standard care | Participants 34126 / 31488; cancer deaths 34 / 64; stage III–IV cases 39 / 82 | FIGO stage II or higher (cervical proxy) | Source values confirmed by two reviewers | None | Confirmed without correction; included | Abstract, PDF p 1: denominators and endpoint definition; Table 4, PDF p 8: mortality and FIGO stage II or higher |
| Telemark Polyp Study I | 8898422 | Invitation to flexible sigmoidoscopy, polypectomy and surveillance / no invitation | Participants 400 / 399; cancer deaths 1 / 1; stage III–IV cases 1 / 2 | Dukes C/D (colorectal mapping) | Source values confirmed by two reviewers | Author-selected earliest-compatible deaths 1/1 and Dukes C/D cases 1/2; the later counts were 1/3 and 1/5 | Included using the author-selected earliest-compatible tuple | Table IV, PDF p 4: colorectal-cancer mortality; text and Table V, PDF p 4: Dukes C/D |
| Randomized Hepatocellular Carcinoma Surveillance Trial | 23750991 | Biannual ultrasonography / annual triple-phase computed tomography; both groups received alpha-fetoprotein testing every 6 months | Participants 83 / 80; cancer deaths 5 / 7; stage III–IV cases 4 / 3 | TNM III/IIIA/IVB | Source values confirmed by two reviewers | None; intervention/control orientation confirmed | Confirmed without correction; included | Figure 1, PDF p 3, and Table 1, PDF p 4: denominators; Table 3, PDF p 4: deaths and stage categories |
| Czech Lung Screening Trial | 2404878 | Semiannual chest radiography plus sputum cytology / control | Participants 3171 / 3174; cancer deaths 85 / 67; stage III–IV cases 53 / 46 | Trial-reported stage III | Source values confirmed by two reviewers | Author-selected deaths 85/67 from the diagnosis cohort and stage-assessment window; later 15-year deaths were 247/216 | Included using the author-selected earliest-compatible tuple | PDF p 1: denominators; Table II, PDF p 4: stage; PDF p 5: lung-cancer mortality |
| LungSEARCH Trial | 31537697 | Low-dose computed tomography surveillance after risk selection / usual care | Participants 785 / 783; cancer deaths 16 / 21; stage III–IV cases 19 / 17 | NSCLC III–IV or extensive-disease SCLC | Source values confirmed by two reviewers | None | Confirmed without correction; included | PDF p 4: denominators; Tables 3 and 4, PDF pp 6–7: stage; PDF p 8: lung-cancer mortality |

**Supplementary Table S6. Complete 49-comparison analytic dataset comprising 41 original Feng et al. comparisons and eight earliest-compatible audit-recovered comparisons used in the updated stage III–IV incidence–mortality sensitivity analysis.**

| **Dataset component** | **Source report or trial family** | **Cancer type** | **Cancer-specific mortality reduction (%)** | **Stage III–IV incidence reduction (%)** | **Mortality significant at P<0.05** | **Stage III–IV incidence significant at P<0.05** |
| --- | --- | --- | --- | --- | --- | --- |
| Original Feng et al. comparison | Miller et al, J Natl Cancer Inst 2000 | Breast | -1.8 | 17.8 | No | No |
| Original Feng et al. comparison | Miller et al, Ann Intern Med 2002 | Breast | 2.8 | -4.4 | No | No |
| Original Feng et al. comparison | Mittra et al, BMJ 2021 | Breast | 14.2 | 17.9 | No | Yes |
| Original Feng et al. comparison | Roberts et al, Lancet 1990 | Breast | 15.6 | 36.1 | No | Yes |
| Original Feng et al. comparison | Andersson et al, BMJ 1988 | Breast | 4.1 | 18.2 | No | No |
| Original Feng et al. comparison | Ramadas et al, Cancer 2023 | Breast | -2.4 | -17.7 | No | No |
| Original Feng et al. comparison | Kronborg et al., Scand J Gastroenterol 1989 | Colorectal | 27.5 | 6.9 | No | No |
| Original Feng et al. comparison | Faivre et al, Gastroenterology 2004 | Colorectal | 16.6 | 14.8 | Yes | No |
| Original Feng et al. comparison | Lindholm, et al, Br J Surg 2008 | Colorectal | 16.0 | 12.0 | Yes | No |
| Original Feng et al. comparison | Hardcastle et al, Lancet 1996 | Colorectal | 14.6 | 8.4 | Yes | No |
| Original Feng et al. comparison | Mandel et al, N Engl J Med 1993 | Colorectal | 33.0 | 24.0 | Yes | Yes |
| Original Feng et al. comparison | Mandel et al, N Engl J Med 1993 | Colorectal | 4.5 | 5.3 | No | No |
| Original Feng et al. comparison | Hoff et al, BMJ 2009 | Colorectal | 27.0 | 10.4 | No | No |
| Original Feng et al. comparison | Bretthauer et al, N Engl J Med 2022 | Colorectal | 8.4 | 19.7 | No | No |
| Original Feng et al. comparison | Niv et al, Gut 2002 | Colorectal | 20.3 | 22.4 | No | No |
| Original Feng et al. comparison | Schoen et al, N Engl J Med 2012 | Colorectal | 26.1 | 29.1 | Yes | Yes |
| Original Feng et al. comparison | Segnan et al, J Natl Cancer Inst 2011 | Colorectal | 21.7 | 26.3 | No | Yes |
| Original Feng et al. comparison | Chen et al, Journal of medical screening 2003 | Liver | -0.7 | 46.5 | No | Yes |
| Original Feng et al. comparison | Zhang et al, J Cancer Res Clin Oncol 2004 | Liver | 40.3 | 47.2 | Yes | Yes |
| Original Feng et al. comparison | Infante et al, Am J Respir Crit Care Med 2009 | Lung | 6.3 | -7.1 | No | No |
| Original Feng et al. comparison | Saghir et al, Thorax 2012 | Lung | -36.4 | -37.5 | No | No |
| Original Feng et al. comparison | Sullivan et al, Eur Respir J 2021 | Lung | 28.8 | 26.3 | No | No |
| Original Feng et al. comparison | Paci et al, Thorax 2017 | Lung | 29.2 | 24.2 | No | No |
| Original Feng et al. comparison | Becker et al, Int J Cancer 2020 | Lung | 27.7 | 43.3 | No | Yes |
| Original Feng et al. comparison | Pastorino et al, Ann Oncol 2019 | Lung | 27.5 | 22.3 | No | No |
| Original Feng et al. comparison | Marcus et al, J Natl Cancer Inst 2000 | Lung | -10.6 | -2.8 | No | No |
| Original Feng et al. comparison | Melamed et al, Chest 1984 | Lung | 7.9 | -6.2 | No | No |
| Original Feng et al. comparison | De Koning et al, N Engl J Med 2020 | Lung | 23.9 | 28.9 | Yes | Yes |
| Original Feng et al. comparison | Aberle et al, N Engl J Med 2011 | Lung | 19.6 | 21.0 | Yes | Yes |
| Original Feng et al. comparison | Oken et al, JAMA 2011 | Lung | 1.4 | 2.4 | No | No |
| Original Feng et al. comparison | Field et al, Lancet Reg Health Eur 2021 | Lung | 34.8 | 56.8 | No | Yes |
| Original Feng et al. comparison | Ji et al, Ann Oncol 2019 | Nasopharyngeal | 31.1 | 22.0 | No | No |
| Original Feng et al. comparison | Ramadas et al, Oral Oncol 2003 | Oral | 8.8 | 10.1 | No | No |
| Original Feng et al. comparison | Buys et al, JAMA 2011 | Ovarian | -18.2 | -19.2 | No | No |
| Original Feng et al. comparison | Jacobs et al, Lancet 2016 | Ovarian | 14.7 | 16.9 | No | Yes |
| Original Feng et al. comparison | Jacobs et al, Lancet 2016 | Ovarian | 11.2 | 9.0 | No | No |
| Original Feng et al. comparison | Jacobs et al, Lancet 1999 | Ovarian | 49.9 | 38.8 | No | No |
| Original Feng et al. comparison | Martin et al, JAMA 2018 | Prostate | 1.7 | 7.7 | No | Yes |
| Original Feng et al. comparison | Schröder et al, N Engl J Med 2009 | Prostate | 19.2 | 6.6 | Yes | No |
| Original Feng et al. comparison | Sandblom et al, Eur Urol 2004 | Prostate | -3.9 | 12.4 | No | No |
| Original Feng et al. comparison | Andriole et al, N Engl J Med 2009 | Prostate | -12.2 | 9.6 | No | No |
| Earliest-compatible audit-recovered addition | Stockholm Mammography Trial (PMID 7579497) | Breast | 49.4 | 16.0 | Yes | No |
| Earliest-compatible audit-recovered addition | Swedish Two-County Trial (PMID 7497137) | Breast | 33.6 | 36.5 | Yes | Yes |
| Earliest-compatible audit-recovered addition | UK Age RCT (PMID 33141657) | Breast | 12.5 | 10.6 | No | No |
| Earliest-compatible audit-recovered addition | Rural India HPV Screening Trial (PMID 19339719) | Cervical | 51.0 | 56.1 | Yes | Yes |
| Earliest-compatible audit-recovered addition | Telemark Polyp Study I (PMID 8898422) | Colorectal | 0.3 | 50.1 | No | No |
| Earliest-compatible audit-recovered addition | Randomized Hepatocellular Carcinoma Surveillance Trial (PMID 23750991) | Liver | 31.2 | −28.5 | No | No |
| Earliest-compatible audit-recovered addition | Czech Lung Screening Trial (PMID 2404878) | Lung | −27.0 | −15.3 | No | No |
| Earliest-compatible audit-recovered addition | LungSEARCH Trial (PMID 31537697) | Lung | 24.0 | −11.5 | No | No |

*Values for the 41 original comparisons reproduce the rounded percentage reductions and statistical-significance classifications reported in Feng et al.’s Supplementary eTable 2. For the eight audit-recovered additions, percentage reduction was calculated as 100 × [1 − (event risk in the intervention group/event risk in the control group)] using the earliest source report containing compatible randomized-arm cancer-specific mortality and stage-category data. Statistical-significance classifications for the additions were calculated using two-sided tests of proportions with continuity correction at α=0.05*

**Supplementary Table S7. Unit definitions for Feng et al. source-review data and the LLM-assisted audit**

| Unit | Count | Level | Role in this manuscript |
| --- | --- | --- | --- |
| Publications included by Feng et al. | 60 | Publication | Source-review publication set used to derive the inherited PMID reference set. |
| Reference-set publications with PMIDs available | 58 | Publication/PMID | Operational reference set for audit-rule agreement among the 1,209 candidate records. |
| Feng et al.’s primary-analysis publications | 39 | Publication | Publications contributing to Feng et al.’s primary stage III-IV, earliest-follow-up analysis. |
| Feng et al. primary paired comparisons | 41 | Intervention-control comparison | Primary framework updated in the sensitivity analysis; multi-arm trials could contribute separate eligible paired comparisons. |
| Feng et al. all-follow-up comparisons | 63 | Follow-up comparison | Broader follow-up-level dataset; not used as the primary update target because it retains multiple follow-up points from the same trial. |
| LLM-prioritized publications absent from the reference set | 33 | Publication/PMID | Publications retained by the 5-of-5 audit rule but absent from the reference set; all underwent human reassessment. |
| Human-confirmed eligible additional publications | 26 | Publication | Publications absent from the reference set and confirmed as eligible after human reassessment. |
| Reconciled unique trials | 18 | Unique trials | Human-confirmed publications grouped by underlying randomized trial to avoid double-counting. |
| Analyzable newly identified comparisons | 8 | Unique-trial records | Reviewer-verified records with compatible randomized-arm endpoint and denominator data added to the 41 record primary framework. |

**Supplementary Table S8. Publications retained and reference-set agreement performance across five-assessment consensus thresholds**

| Consensus rule | Total PDF-available publications meeting rule | Reference-set publications recovered | Non-reference publications requiring reassessment | Potential reduction in non-reference reassessment workload, n/938 (%) |
| --- | --- | --- | --- | --- |
| Any-of-5 include | 118 | 58 | 60 | 878 (93.6%) |
| 2-of-5 include | 108 | 58 | 50 | 888 (94.7%) |
| Majority include | 106 | 58 | 48 | 890 (94.9%) |
| 4-of-5 include | 100 | 58 | 42 | 896 (95.5%) |
| 5-of-5 include | 91 | 58 | 33 | 905 (96.5%) |

**Supplementary Appendix S1. Verbatim endpoint-oriented prompt and output categories used for multimodal PDF review.**

**1. ROLE & OBJECTIVE**

You are a world-renowned researcher specializing in cancer prevention and an expert in conducting rigorous, error-free systematic reviews. Your task is to conduct a detailed full-text review to make a definitive **INCLUDE** or **EXCLUDE** decision to be published in a top-tier medical journal.

The review's objective is to **determine whether the incidence of late-stage cancer is a suitable alternative endpoint to cancer-specific mortality in cancer screening RCTs.**

Your primary goal is **INCLUSION**. You must exhaust all possibilities to find or derive the required data before recommending exclusion. Assume the original authors were able to include this paper and your job is to figure out how they did it.

**2. CONTEXT: THE PAPER TO ANALYZE**

The full text of the research paper you must analyze is contained in the attached file:

**{FULL MANUSCRIPT CONTENT}**

**3. SCREENING CRITERIA (PICO-S)**

A study is eligible only if it meets **ALL** of the following criteria. If the manuscript states clearly fails to meet **ANY** of these, it must be excluded.

- **[L] Language:** The study's primary text must be English

- **[P] Population:** The trial's participants must be from a general or at-risk **asymptomatic population** eligible for screening.

- **Exclude if:** The study exclusively enrolls participants who already have a confirmed cancer diagnosis or are being evaluated for symptoms.

- **[I] Intervention:** The intervention must be a **cancer screening test** or strategy (e.g., imaging, endoscopy, biomarker test).

- **Exclude if:** The intervention is purely therapeutic (e.g., drug trial), preventive (e.g., vaccine, chemoprevention), or a diagnostic workup for symptomatic patients.

- **[C] Comparator:** The trial must have a distinct comparator group, such as **usual care, no screening, or an alternative screening modality.**

- **Exclude if:** It is a single-arm study or if the comparison group receives the exact same screening intervention under the same protocol (i.e., not a true comparison).

- **[O] Outcomes:** The study must report, or be reasonably expected to report, data on **both cancer-specific mortality AND cancer stage.**

- **Heuristic:** If an abstract for an RCT on cancer screening mentions mortality as an endpoint, assume stage data is likely available in the full text and do not exclude. Exclude only if the abstract *explicitly states* that one or both outcomes were not measured.

- **[S] Study Design:** The study must be a **pre-specified long-term follow-up of a Randomized Controlled Trial (RCT)** that used either individual or cluster randomization. The primary language must be **English**.

- **Ineligible Study Designs (Must Exclude)**:

- Systematic reviews, meta-analyses, and narrative reviews.

- Observational studies (cohort, case-control, cross-sectional).

- Non-randomized or quasi-experimental trials.

- Study protocols, commentaries, editorials, or letters without original trial data.

- **True Secondary analyses:** These studies use data from an existing RCT to answer a *new* or *different* research question that was not the trial's original primary objective (Please use this with caution and rare. We would rather you read the full manuscript to make the decision).

- **Do NOT Exclude (Potentially Eligible):**

- **Long-term follow-up reports:** Publications that provide updated results on the trial's original primary endpoints (mortality, stage) after a longer period. These are crucial and must be included.

**4. REQUIRED DATA POINTS**

For a study to be included, you must extract the absolute numbers for the following four endpoints:

1. **Cancer-Specific Deaths (Intervention Group)**

2. **Cancer-Specific Deaths (Control/Comparison Group)**

3. **Late-Stage (III-IV) Cancer Cases (Intervention Group)**

4. **Late-Stage (III-IV) Cancer Cases (Control/Comparison Group)**

**5. STAGING SYSTEM CLASSIFICATION GUIDE**

This is the most critical part of your task. Studies rarely use a simple "Stage I-II vs III-IV" format. The original review authors used the following specific rules to convert various staging systems. **You MUST apply these rules to interpret the data.

Your primary goal is to find the absolute number of cases that fit the **"Late-Stage"** definition for each cancer type according to these specific rules. You can apply these rules to interpret the data from the manuscript.

| Cancer Type | Original Stage System Reported in Trial | How to Map to Late-Stage (III-IV) |

| **Breast Cancer** | Tumor size | **Late-Stage = Tumor ≥20 mm** |

| **Colorectal Cancer** | Dukes staging | **Late-Stage = Dukes C + Dukes D** |

| | Localized and advanced | **Late-Stage = "Advanced" cases** |

| **Lung Cancer** | Early/late and resection | **Late-Stage = "Late stage unresected" cases** |

| **Prostate Cancer** | **Risk Groups** (TNM + Gleason + PSA) | **Late-Stage = "High risk" cases + "Advanced" cases** <br>*(Note: "Low risk" and "Intermediate risk" are considered early-stage and should be excluded from the late-stage count.)* |

| | **Gleason score** only | **Late-Stage = Gleason 8-10 cases + "Metastatic" cases** <br>*(Note: Gleason 2-6 and Gleason 7 are considered early/intermediate stage and should be excluded from the late-stage count. This is a critical distinction.)* |

| | **TNM-based classification** | **Late-Stage = "Advanced tumours,"** defined as cases with T3–4, or N1, or M1. <br>*(Note: "Localised tumours," defined as T1–2 AND N0/NX AND M0, are early-stage.)* |

| **Any Cancer** | Standard TNM or UICC Staging or AJCC Cancer Staging Manual or any manual that provides cancer stage information | **Late-Stage = Stage III cases + Stage IV cases**. If the paper provides a direct breakdown by Roman numeral stage (I, II, III, IV), this is the preferred method. |

**Your Mandate:** During the extraction process, you must actively search for these specific original staging systems within the text and tables of the full manuscript. If you find one, apply the corresponding rule from this guide to calculate the number of late-stage cases. Do not exclude a paper if the staging data is present in one of these formats.

**6. SCREENING ANALYSIS PROTOCOL (MANDATORY CHAIN-OF-THOUGHT)**

You must follow these steps in your reasoning before providing the final JSON output.

1. **Initial PICO-S Analysis:** Briefly summarize the study's Population, Intervention, Comparator, Outcomes, and Study Design as described in the abstract.

2. **Criterion-by-Criterion Evaluation:** Assess the abstract against each of the 5 screening criteria. For each criterion, state whether it is Met, Not Met, or Uncertain from abstract and provide a one-sentence rationale.

- **Population:**

- **Intervention:**

- **Comparator:**

- **Outcomes:**

- **Study Design:**

Once confirm the study meets the inclusion criterion, please further verify whether the data can be extracted:

3. **DATA EXTRACTION** :

* **Step 1: Extract Cancer-Specific Mortality Data**: Search the full text (especially tables and Results section) for the absolute number of cancer-specific deaths in each study arm. If found, state the numbers and the source (e.g., "Table 3"). If not found, state that mortality data is missing.

* **Step 3: Extract Late-Stage (Stage III-IV) Cancer Case Data.**: This requires a hierarchical search. **Do not stop until you have tried all sub-steps.**: **3a. Direct Search:** Look for tables or text that explicitly list the number of cases by Stage I, II, III, and IV. If you find them, sum the numbers for Stage III + IV for each arm. **3b. Mapped Search (Apply the Guide):** If direct stages are not available, use the **Staging System Classification Guide** from section 3. Search the paper for alternative systems (e.g., Dukes staging, Gleason scores, tumor size, risk groups). Apply the rules to calculate the number of late-stage cases. For example, if you find the number of patients with Gleason 8-10, count them as late-stage. **3c. Proxy Search:** Look for the absolute number of cases described with proxy terms like "advanced cancer," "metastatic disease," "late-stage disease," "unresectable disease." If found, use these numbers. **3d. Calculation Check:** If the paper only provides incidence *rates* (e.g., "1.6 per 1000 person-years") and the total *person-years* or *total participants*, state this. Do not exclude; identify the components needed for a human to perform the calculation.

* **Step 4: Check for Supplemental Material.** Scan the paper (Methods, Results, table footnotes) for any mention of "Supplementary Material," "Appendix," or "online supplement." If the required data is stated to be in a supplement, this is a critical finding.

4. **Synthesize and Render Final Verdict.** Based on your investigation, choose one of the three verdicts and provide the reasoning.

- **INCLUDE**: You successfully found or derived all four required data points from the main text using the protocol.

- **INCLUDE_DATA_IN_SUPPLEMENT**: The paper is eligible, but one or more essential data points are located exclusively in the supplementary materials, which are not currently available to you.

- **EXCLUDE**: After applying all steps of the protocol (including all mapping and proxy rules), the data is definitively and irretrievably missing from the main text, and no mention is made of it being in a supplement. This should be a rare verdict.

**6. FINAL OUTPUT FORMAT (MANDATORY)**

````
Your entire response must be a single, valid JSON object.

Generated json

```
{
  "verdict": "INCLUDE | INCLUDE_DATA_IN_SUPPLEMENT | EXCLUDE",
  "reasoning": "A concise summary of your extraction process and the final justification for your verdict. If EXCLUDE, state exactly which data point is irretrievably missing and why the mapping/proxy rules could not be applied.",
  "evidence_quote": "Direct quote(s) from the paper that support your data extraction or your verdict.",
  "extracted_data": {
    "mortality_intervention": "Number or 'Not Found'",
    "mortality_control": "Number or 'Not Found'",
    "late_stage_intervention": "Number, 'Calculation Required', or 'Not Found'",
    "late_stage_control": "Number, 'Calculation Required', or 'Not Found'",
    "source_mortality": "e.g., Table 2, Page 5",
    "source_stage": "e.g., Figure 1 (interpreted using Gleason score mapping), Page 6"
  }
}
```
````

**Supplementary Appendix S2. Technical implementation details for the LLM-assisted full-text audit.**

Gemini 3.1 Pro Preview was selected because it supported native PDF input, a 1,048,576-token context window, multimodal document understanding, structured output, and API-level parameter control suitable for long clinical reports containing tables and figures (reference 19 in the main manuscript). The same prompt and settings were used throughout. Repeated assessment was intended to characterize consistency within one model configuration rather than agreement among independent models or external validation; a comparable repeated-assessment approach has been used in LLM-assisted screening (reference 20 in the main manuscript).

| **Implementation element** | **Specification** |
| --- | --- |
| **Model and provider route** | Google Gemini 3.1 Pro Preview through the OpenRouter application programming interface. |
| **Document input** | PDFs were base64-encoded and submitted through OpenRouter's native PDF route without local text-layer extraction, optical character recognition, or layout reconstruction. |
| **Request context** | Each assessment was a separate request containing the fixed system prompt, fixed user instruction, and one PDF, without prior conversational messages. |
| **Repetitions and date** | Five separate assessments per PDF; assessments were completed in April 2026. |
| **Temperature** | Temperature was set to 1.0 in accordance with model-specific guidance. |
| **Other sampling controls** | Maximum output tokens, top-p, top-k, presence penalties, frequency penalties, and other sampling controls were not specified; OpenRouter defaults in effect at inference were used and exact default values were not archived. |
| **Ordering and seed** | PDF order and repetition order were not randomized, and no randomization seed was used. |
| **Network-failure handling** | Network or HTTP failures were retried for up to five total attempts, with a 300-second timeout per attempt and exponential backoff of 1, 2, 4, and 8 seconds. |
| **Content-error handling** | Responses with absent or unparseable model content were recorded as errors and were not automatically resubmitted. |
| **Successful output** | A decision category, reasoning or evidence, source locations, and provisional intervention- and control-arm mortality and late-stage counts. |
| **Assessment yield** | Among 4,980 assessments, 4,949 returned a recognized verdict and 31 (0.6%) returned an error, affecting 29 publications; all 31 errors involved absent model content. |
| **Interpretation of repetitions** | The five repetitions assessed consistency under one fixed configuration; they were not treated as independent validators or as external validation. |
